# Transmission patterns of *Candida auris* among regional healthcare facilities revealed by genomic epidemiology and patient transfer data

**DOI:** 10.1101/2025.06.25.25330301

**Authors:** Hannah J. Barbian, Kelly A. Walblay, Alyse Kittner, Christy Zelinski, Erin Newcomer, Hira Adil, Stefan J. Green, Ann Valley, Do Young Kim, Stephanie R. Black, Mary K. Hayden

## Abstract

*Candida auris* emerged in Chicago, IL, USA in 2016 and has since become endemic. We used whole genome sequencing (WGS) of 494 isolates, epidemiologic metadata, and patient transfer data to describe the transmission of *C. auris* among Chicago healthcare facilities between 2016 - 2021. 99% of isolates formed a single clade IV phylogenetic lineage, suggesting a single introduction. Isolates were grouped into genomic clusters based on phylogenetic clustering and relatedness. Isolates from 14 facilities were included in 77 genomic clusters with evidence of inter- and intra-facility transmission; 62% of isolates included in genomic clusters clustered with another from the same facility. Patient transfer data corroborated transmission among facilities in large genomic clusters and patient transfers between facilities correlated with more similar genomes. 30% of individuals who had serial samples collected over time had isolates collected during different facility stays. Most of these individuals carried closely related isolates over time, although 25% carried isolates whose genomes diverged. Integrating WGS data and patient metadata to describe inter- and intra-facility clusters revealed regional transmission patterns within and between healthcare facilities that could be used to guide targeted interventions to prevent transmission. Given that patient transfer data were concordant with WGS analysis, they may serve as a predictive tool for risk of inter-facility transmission.

## INTRODUCTION

*Candida auris* emerged in Chicago in 2016 when two cases were identified in a single healthcare facility (*1*). Since then, cases have increased citywide and *C. auris* has become endemic (*2*). *C. auris* poses a high risk to patients given its ability to cause severe illness, persist in the environment, and develop resistance to antifungal agents (*3–8*). *C. auris* has been documented to cause outbreaks in healthcare settings, especially post-acute care settings such as long-term acute care hospitals (LTACHs) and ventilator-capable skilled nursing facilities (vSNFs), and can affect entire patient wards, resulting in prevalence as high as 71% (*8–13*).

Prolonged colonization and poor implementation of infection control practices such as hand hygiene and environmental cleaning in long-term care settings increases the risk of <u>intra-facility</u> <u>transmission</u> (*14*). Acute care facilities that are nodes in transfer networks with long-term care facilities may be at risk for admitting patients with *C. auris*, leading to <u>inter-facility transmission</u>. The Chicago Department of Public Health (CDPH) has prioritized *C. auris* prevention efforts and conducts point prevalence surveys (PPS) at facilities admitting patients from high burden post-acute care settings as informed by patient transfer data.

The global emergence of *C. auris* has been monitored using whole genome sequencing (WGS) and genomic epidemiology. Genomic analysis has revealed that six clades of *C. auris* (clades I-VI) emerged near-simultaneously on multiple continents and subsequently spread to other locations worldwide (*15–18*). Once introduced to a new location, *C. auris* may spread rapidly, as evidenced by identification of many new cases and closely related isolates within geographic areas (*19–22*). However, independent introductions of multiple *C. auris* clades to a single geographic location have also been described (*23, 24*). While genomic diversity is often limited during early regional spread of *C. auris*, isolates collected from the same individual and from epidemiologically-linked cases are generally more genetically similar than unrelated regional isolates (*8, 11, 21*). This indicates that genomic analysis may be useful for identifying patterns of transmission in regional *C. auris* isolates, especially as greater genetic diversity evolves amongst local isolates over time. *C. auris* genomic studies have been reported at the broad (e.g., national) scale and at the individual institutional scale; however, these investigations have lacked integration of patient metadata or have described limited outbreaks. Genomic epidemiology has yet to be applied to understand *C. auris* dissemination at a regional level. Further, bioinformatic identification of *C. auris* clusters has yet to be developed for unbiased selection of related isolates in large genomic datasets.

Here, CDPH collaborated with the Wisconsin State Laboratory of Hygiene (WSLH) and the Chicago-based Regional Innovative Public Health Laboratory (RIPHL) to integrate WGS results, epidemiologic metadata, and patient transfer information to describe the evolution and transmission of *C. auris* in Chicago healthcare facilities from the introduction of *C. auris* in 2016 through December 2021 and establishment of endemicity.

## METHODS

### *C. auris* case reporting

*C. auris* has been reportable to the Illinois Department of Public Health (IDPH) since 2018. Case definitions for reporting follow the Centers for Disease Control and Prevention (CDC) National Notifiable Diseases Surveillance System definition: A clinical case is defined as *C. auris* identified from a specimen obtained during clinical care, whereas a screening case is defined as *C. auris* identified from a swab collected for the purpose of screening for *C. auris* colonization (*30, 31*). The first screening case and subsequent clinical case (if available) for each person is reported to the IDPH eXtensively Drug Resistant Organism (XDRO) Registry (*32*), a bidirectional registry of individuals with a history of *C. auris* (and other multidrug-resistant organisms) that facilitates public health department tracking of case counts and improves inter-facility communication by allowing facilities to query the registry for new admissions to ensure patients are placed on appropriate transmission-based precautions (*33*).

### Active surveillance

In addition to the passive reporting by healthcare facilities, CDPH conducts active surveillance for *C. auris* colonization in selected healthcare facilities. During August 2018- December 2021, <u>biannual PPSs</u> were conducted in four vSNFs and four LTACHs in Chicago. <u>Response PPSs</u> were conducted in additional healthcare facilities when cases were noted to have increased or when *C. auris* was identified in a facility with no prior cases reported. A third category of <u>prevention-based PPSs</u> were conducted by CDPH for ACHs to assess burden of *C. auris* in acute care settings, specifically intensive care units. Response and prevention PPSs were conducted as health department resources allowed.

For all PPSs facilitated by CDPH, a composite bilateral axilla/groin swab was collected from residents and specimens were shipped to WSLH for *C. auris* PCR and culture testing (*34*). Additionally, during 2019-2021, CDPH facilitated *C. auris* admission screening at selected LTACHs using the same sample collection and testing protocol. All *C. auris* isolates from CDPH-conducted PPS and admission screenings between 2018-2021 were stored frozen at -80°C at WSLH.

### Whole genome sequencing

Available stored *C. auris* isolates collected August 2018-December 2021 were transported to RIPHL, which is a public-academic partnership between CDPH and Rush University Medical Center to provide CDPH with the capacity for advanced molecular detection and surveillance of pathogens of public health interest. Following nucleic acid extraction using the Maxwell RSC Cultured Cells DNA Kit and Maxwell automated extraction system (Promega), sequencing libraries were prepared using 1 ng of DNA extract and the Nextera XT DNA Library Preparation Kit (Illumina). Genome libraries were barcoded using IDT for Illumina DNA/RNA UD Indexes (Illumina) and balanced using a small-scale sequencing run of an equivolume pool on an Illumina iSeq. Pooled libraries were sequenced at a depth of approximately 10 million clusters per sample on an Illumina NovaSeq6000 or NovaSeqX sequencer, employing paired-end 2x150 base sequencing. Raw sequences have been submitted to NCBI SRA repository under the BioProject PRJNA904373 (Supplementary Table 1).

### Bioinformatic analysis

Sequencing data were analyzed using the MycoSNP-nf pipeline (v.1.4) using the Clade IV reference B11243 (GCA_003014415.1) on Terra.bio (*23, 35*). Isolates with estimated coverage depth <25 were excluded from downstream analysis. *C. auris* clade assignment was determined by assessing phylogenetic clustering of isolates with Clade I – IV reference sequences (Supplementary Table 1). MycoSNP-nf multi-fasta files were used to build a neighbor-joining phylogenetic tree including all sequences using MEGA 11 with 100 bootstrap replicates as previously described (*21, 36, 37*). All Illinois *C. auris* sequences available in NCBI SRA were also included (Supplementary Table 1). MycoSNP-nf distance matrices were used to evaluate SNP differences between isolates. Clusters including two or more isolates were identified using ClusterPicker (*38*) with clade bootstrap support >80% and maximum genetic distance 0.3%. MycoSNP-nf-derived multi-fasta files containing all SNP differences to the reference and within selected sequences were used for ClusterPicker input. For the input dataset, the multi-fasta sequence alignment length was 2,403 nucleotide positions, and a genetic distance of 0.3% corresponded to ∼7 SNPs. This maximum genetic distance was selected based on examining the fraction of isolate pairs that are intra-facility at pairwise SNP distances and selecting the SNP distance where there is a decline, as reported previously, as well as the decline in the frequency of pairwise SNP distances between isolates collected from the same individual (Supplementary Fig. 1, Fig. 2c). This SNP distance approximately corresponds to SNPs expected to accumulate in a *C. auris* genome over one year based on the estimated mutation rate (*8, 26*). A sensitivity analysis using a genetic distance of 0.2, 0.3, 0.4 and 0.5% was performed; the selected 0.3% genetic distance mostly closely aligned with visual identification of clustering sequences. “Intra-facility” pairs were defined as two isolates from the same facility and “inter-facility” pairs were defined as two isolates from different facilities at the time of isolate collection. If facility was unknown, e.g. for contextual isolates, intra/inter-facility status was not assigned. Small genomic clusters were defined as those with fewer than five isolates and large genomic clusters as five or more.

### Patient transfer data

Patient transfer information was derived from 2020 Centers for Medicare and Medicaid Services (CMS) fee for service beneficiary claims data linked to the CMS Minimum Data Set by Medicare beneficiary identifier. The dataset comprises information from Illinois medical facilities including LTACHs, ACHs and SNFs, but is not representative of an all-payer network.

Direct patient transfers were analyzed; the number of direct transfers between two facilities is enumerated by the number of discharges from one facility admitted directly to another facility within one day. Patient transfer data were not available from all included years. Thus, 2020 patient transfer data was selected as the median collection date for included isolates was January 15, 2020. Network visualizations were constructed for all facilities with sequenced isolates using the igraph package in R with algorithm=auto. These networks were used to visualize potential transmission of *C. auris* among facilities. Patient transfers less than 10 were reported as “9” as required by the CDC data use agreement with CMS. Patient flow analysis was performed on sites with more than 10 transfers using the R package regentrans and plotted using ggplot and the geom_smooth lm method (*26*).

### Statistical analysis

Statistical comparisons for SNP distances were performed using Mann-Whitney test with a two-tailed p-value and Prism v9 software (GraphPad). Correlations were visualized using linear regression and statistical assessment was performed using Spearman’s rank correlation coefficient with a two-tailed p-value (α = 0.05) in Prism v9 software (GraphPad).

## RESULTS

### Chicago *C. auris* surveillance 2018-2021

Between August 1, 2018, and December 31, 2021, 896 new *C. auris* cases (344 clinical and 552 screening) were reported in Chicago (Fig. 1). During this period, CDPH conducted 71 PPSs (41 routine, 13 response, 17 prevention, defined in methods) and facilitated admission screening at three LTACHs for varying time periods (3-10 months). *C. auris* isolates from 496 positive cultures tested at WSLH were available for sequencing and 494 (99.6%) had sufficient genome quality for phylogenetic analysis. These isolates were collected from 420 individuals: One isolate was collected from each of 355 individuals and 139 isolates were collected sequentially from 65 individuals (2-4 isolates). In total, isolates from these individuals were recovered from 19 Chicago metropolitan area facilities (four LTACHs, four vSNFs, two inpatient rehabilitation facilities (IRFs), one skilled nursing facility (SNF), ten acute care hospitals (ACHs)).

**Fig. 1.**
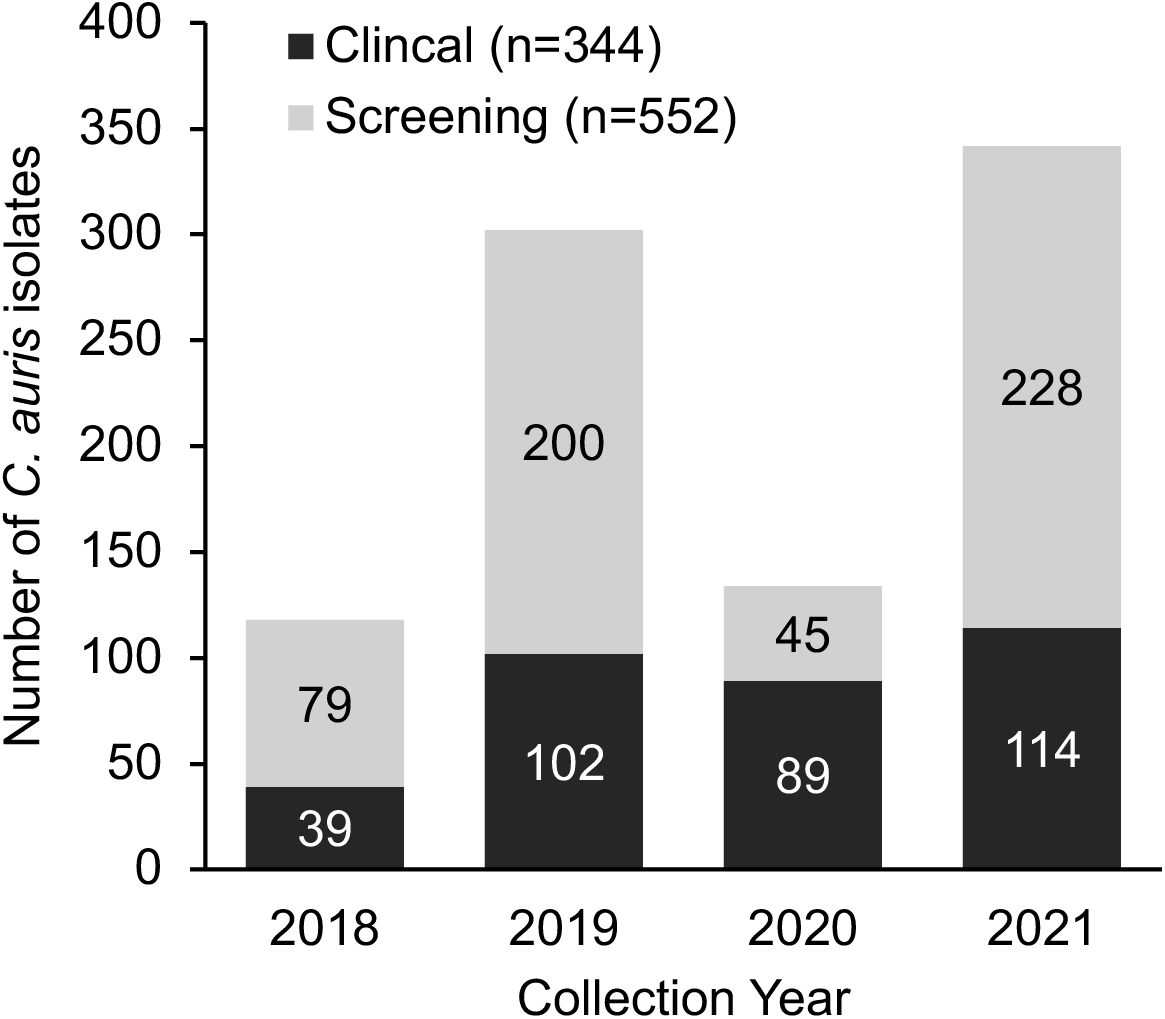
Chicago *C. auris* cases by specimen type, August 1, 2018-December 31, 2021. New screening cases (grey) and clinical infections (black) indicated in bar color. A person may be counted twice, as both a screening and clinical case.

### Phylogenetic and SNP analysis

Of the 494 Chicago *C. auris* isolates sequenced, 491 (99.4%) belonged to clade IV. These isolates were subjected to phylogenetic analysis together with 82 contextual clade IV isolates. These contextual isolates included 15 Chicago-area genomes collected 2018-2021 and sequenced as part of other analyses and 67 publicly available genomes from Illinois collected 2016-2021. All Illinois sequences clustered together with a single common ancestor with early sequences showing very little diversity, suggesting clade IV emerged in Chicago following a single introduction event (Fig. 2a; an interactive version of this tree is available at https://microreact.org/project/gV9gnmBgz18n8fjvyjxZtG). *C. auris* diversity accumulated over time, with mean pairwise SNP differences increasing from 4.3 in 2016 to 30.7 in 2021 (Fig. 2b). Many isolates were minimally diverged from the Illinois ancestor, but phylogenetic clades of related isolates developed over time. These mostly contained only a few isolates; only 12 clades contained ≥10 isolates and 3 contained ≥25 isolates. Isolates did not seem to cluster based on collection date or facility. Rather, genomes from isolates identified in different facilities appear interspersed throughout the tree, suggesting substantial strain sharing among regional facilities (Fig. 2a). However, some groups of related isolates from the same facility were also observed.

**Fig. 2.**
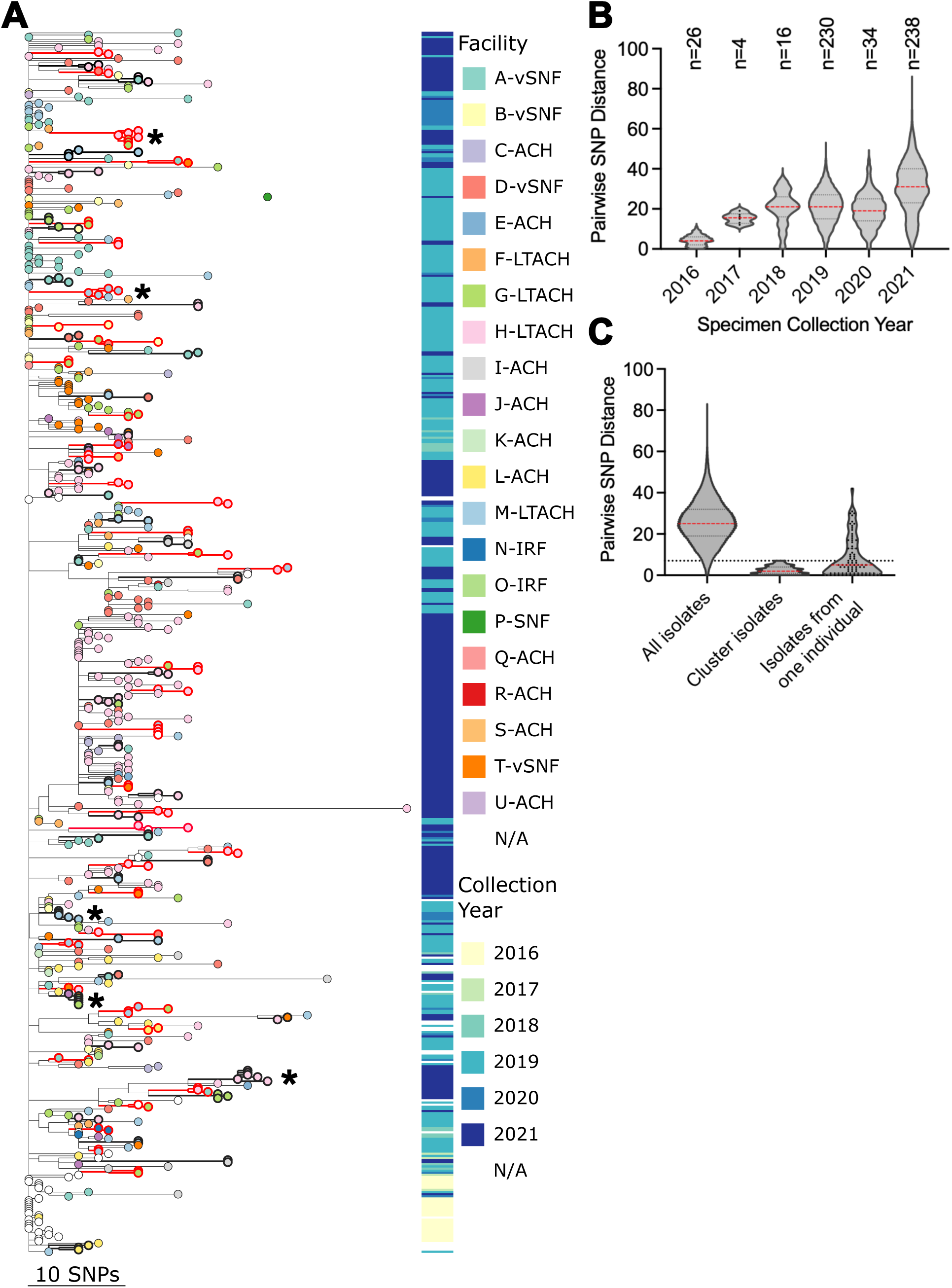
Phylogenetic and SNP analysis of Chicago clade IV *C. auris* isolates. (**A**) Phylogenetic tree of Illinois sequences with tree tips colored by isolate facility. Metadata block to the right of tree tips represents isolate collection year. Isolates included in clusters and branches leading to identified clusters are bolded in alternating black and red for clarity. Asterisks indicate clusters with 5 or more isolates which were selected for subtree visualization in Fig. 3. Scale bar indicates SNP distance. ACH: acute care hospital; LTACH: long-term acute care hospital; IRF: inpatient rehabilitation facility; vSNF: ventilator-capable skilled nursing facility; SNF: skilled nursing facility. (**B – C**) Violin plots of pairwise SNP distances with dashed lines showing median (red) and 25^th^ and 75^th^ quartiles (black); individual data points shown when <100. Isolate numbers shown above plots in b. Dashed horizontal line in panel c indicates the maximum SNP distance of 7 used for cluster selection.

Three isolates (0.6%) from samples collected in 2021 were identified as clade III, representing the first non-clade IV isolates identified in the state. The clade III isolates were closely related (4-8 SNP differences) and collected from the same facility and on the same date from unique individuals, suggesting introduction from a common source and spread within the facility. Thus, while clade IV is the predominant *C. auris* clade in Illinois, other clades including clade III may have been introduced with limited spread or circulating at low frequency.

### Genomic cluster analysis

Given the close genetic relationships among Chicago *C. auris* isolates, especially in the few years following introduction of *C. auris* to Chicago, putative *C. auris* clusters were identified conservatively from these 573 Illinois genomes by phylogenetic clustering (monophyletic clades with >80% bootstrap support) and genetic distance (maximum of 7 SNPs between all isolates in the clade). The phylogenetic clustering threshold was implemented to exclude early isolates containing little genetic diversity. Maximum genetic distance between all isolates in the cluster was implemented to identify very closely related isolates and recent transmission events. Lineages persisting and diversifying over an extended time may be excluded by this conservative method.

A total of 77 genomic clusters were identified with 196 (34%) of 573 sequences included in a cluster (Fig. 2a). Of these, most contained few isolates (mean 2.5 isolates per cluster, range: 2-7 isolates) and were closely related (mean 2.4 pairwise SNP distance within clusters) with strong phylogenetic support (mean bootstrap 95.7%) (Fig. 3a-c). Collection dates for clustering isolates ranged from September 2018 – December 2021, with median collection date difference of 56 days (range 0 - 636). The median pairwise collection date difference between all Illinois clade IV isolates was higher at 447 days (p<0.0001, Mann-Whitney test), indicating that clustering isolates were more likely to be collected closer in time. Twenty clusters (26%) contained isolates collected on the same date and from the same facility, i.e. from a single PPS.

**Fig. 3.**
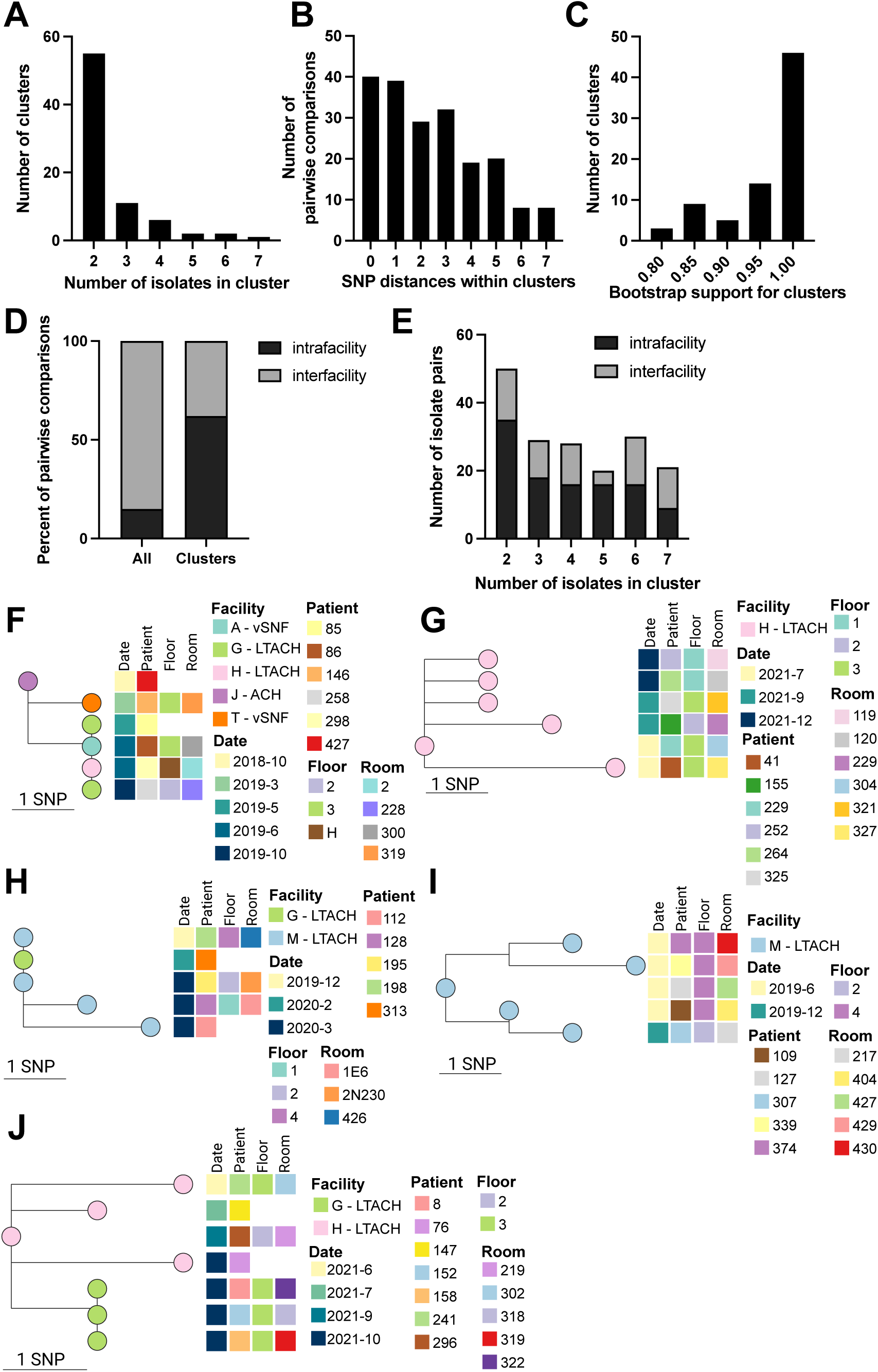
Illinois *C. auris* cluster analysis. (**A-E**) Histograms showing the number of isolates in each cluster (**A**), the pairwise SNP distances contained in each cluster (**B**), bootstrap support for each cluster (**C**) intra/inter-facility relationship between pairwise comparisons of isolates with facility metadata (**D**), and the intra/inter-facility relationship between pairwise comparisons with facility metadata within clusters containing various numbers of isolates (**E**). (**F-J**) Subtrees showing clusters with 5 or more isolates. Subtree tips are colored by isolate facility and relevant isolate/patient metadata are located to the right of tree tips. Scale bar indicates SNPs. ACH: acute care hospital; LTACH: long-term acute care hospital; IRF: inpatient rehabilitation facility; vSNF: -capable skilled nursing facility; SNF: skilled nursing facility.

Fourteen (67%) of 21 facilities with sequenced *C. auris* isolates were included in at least one genomic cluster. All 14 facilities contributed multiple isolates to at least one cluster, with 55 (71%) clusters containing at least two isolates from the same facility (intra-facility clustering). In contrast, 28 (36%) clusters contained isolates from different facilities (inter-facility clustering), and these clusters included 11 facilities.

When comparing isolates pairwise within clusters, intra-facility pairs represented 62% of pairwise comparisons with known facility metadata, considerably increased from 13% of pairwise comparisons between all Illinois clade IV isolates (Fig. 3d). Inter-facility pairs were found less frequently within clusters at 38% of pairwise comparisons, compared to 71% of pairs among all Illinois clade IV isolates (Fig. 3d). This suggests that closely related isolates are more likely to be collected from the same facility. Intra-facility and inter-facility pairs were present in similar proportions in both small and large clusters (Fig. 3e), indicating that intra- or inter-facility transmission did not seem to be associated with cluster size.

Five large genomic clusters, each containing 5-7 isolates collected from unique individuals and hereinafter referred to as Clusters f-j, were identified and epidemiologic metadata were integrated to facilitate interpretation and investigation of isolate linkage (Fig. 3f- j). Cluster f included six isolates collected October 2018 - October 2019 at five facilities, with one isolate collected at the time of patient admission to LTACH G (May 2019). Genomic analysis showed that four of these isolates were identical, providing evidence for strain sharing between multiple facilities. One isolate was collected at LTACH G 5 months later (October 2019), suggesting intra-facility transmission. Cluster g included six isolates collected from three different wards of one LTACH during three separate PPSs spanning five months. The relatedness of these isolates from the same facility over a short timeframe suggests intra-facility transmission. Cluster h included four isolates collected from patients on multiple floors of LTACH M over 4 months, and one isolate from LTACH G identified upon admission during the same period. Three isolate genomes were identical: two collected three months apart at LTACH M and the isolate from the specimen collected upon admission at the LTACH G suggesting strain sharing within and between facilities. Cluster i included four isolates collected from one floor of a LTACH during a PPS and one collected on a different floor during a PPS 6 months later, suggesting strain sharing within the facility. Cluster j included four isolates from LTACH H collected in October 2021 and three identical isolates from LTACH G collected between June and October 2021, suggesting that both inter-facility and intra-facility strain sharing may have occurred. Thus, isolate metadata, i.e., date and/or location of collection, and epidemiologic investigation supported the linkage of isolates included in clusters that were identified by genomic analysis.

### Patient transfer analysis

Patient transfer data revealed that facilities with sequenced *C. auris* isolates are highly inter-connected by patient transfers (Fig. 4a). A few groups of facilities had more frequent transfers, while most connections represented a small number of patient transfers (<10 in a year). Three of five large genomic clusters contained genomic evidence of inter-facility transfers (Fig. 3f, h, j) and patient transfer data corroborated genetic linkages in two of these clusters. In cluster f, the earliest and most genetically ancestral isolate in this cluster was collected from ACH J. Patients were transferred from ACH J to vSNF T, LTACH H and LTACH G, with further transfers from LTACH G to vSNF A (Fig. 4b), supporting the possibility of *C. auris* transfer between these facilities. LTACHs G and H included in cluster j were also linked in the patient transfer data. There were no recorded transfers between LTACHs G and M included in cluster h, indicating potential intermediary transmissions that were not sampled. Patients are not usually admitted directly from the community to an LTACH but rather they are usually transferred from an ACH. According to patient transfer data, LTACHs G and M both received patients from ACH I, K, L, Q, R, S, and U, one or more of which may have served as intermediary facilities where occult transmission of *C. auris* occurred.

**Fig. 4.**
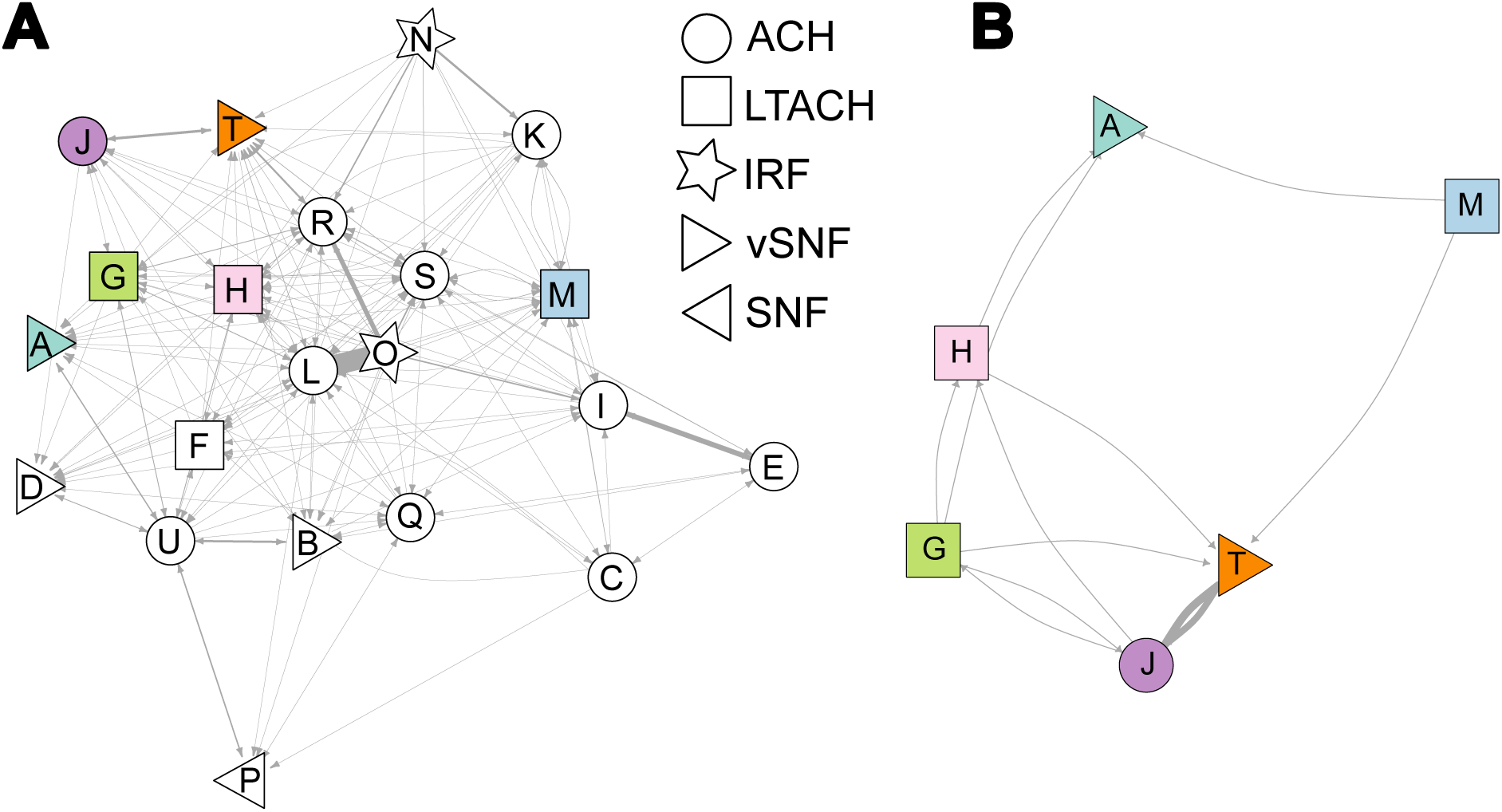
Patient transfer networks for facilities with sequenced *C. auris*. Network analysis of direct patient transfers (connections) for facilities (nodes) from (**A**) all sequenced isolates and (**B**) facilities included in large clusters corresponding to Fig. 3f-j. Facilities included in large clusters are colored according to Fig. 3. Connecting lines are weighted by the number of transfers, with the finest lines indicating fewer than 10 transfers. Vertex shape indicates facility type: ACH: acute care hospital; LTACH: long-term acute care hospital; IRF: inpatient rehabilitation facility; vSNF: ventilator-capable skilled nursing facility; SNF: skilled nursing facility.

To investigate whether facilities with more frequent patient transfers also had more closely related *C. auris* genomes overall, we calculated bi-directional patient flow between facilities with *C. auris* WGS data and >10 transfers and compared this with measures of *C. auris* genomic relatedness (*25, 26*). There was a small positive association between patient flow and the number of closely related *C. auris* pairs (genomes with ≤ 3 SNP differences, selected based on the median within genomic cluster SNP distance), meaning that facilities with more patient transfers had more closely related *C. auris* genome pairs (Fig. 5a, Spearman r=0.23, p<0.0001). There was a weak but significant negative association with patient flow and minimum pairwise SNP distance, indicating that facilities with more transfers contained slightly more closely related genomes (Fig. 5b, Spearman r=-0.13, p=0.017).

**Fig. 5.**
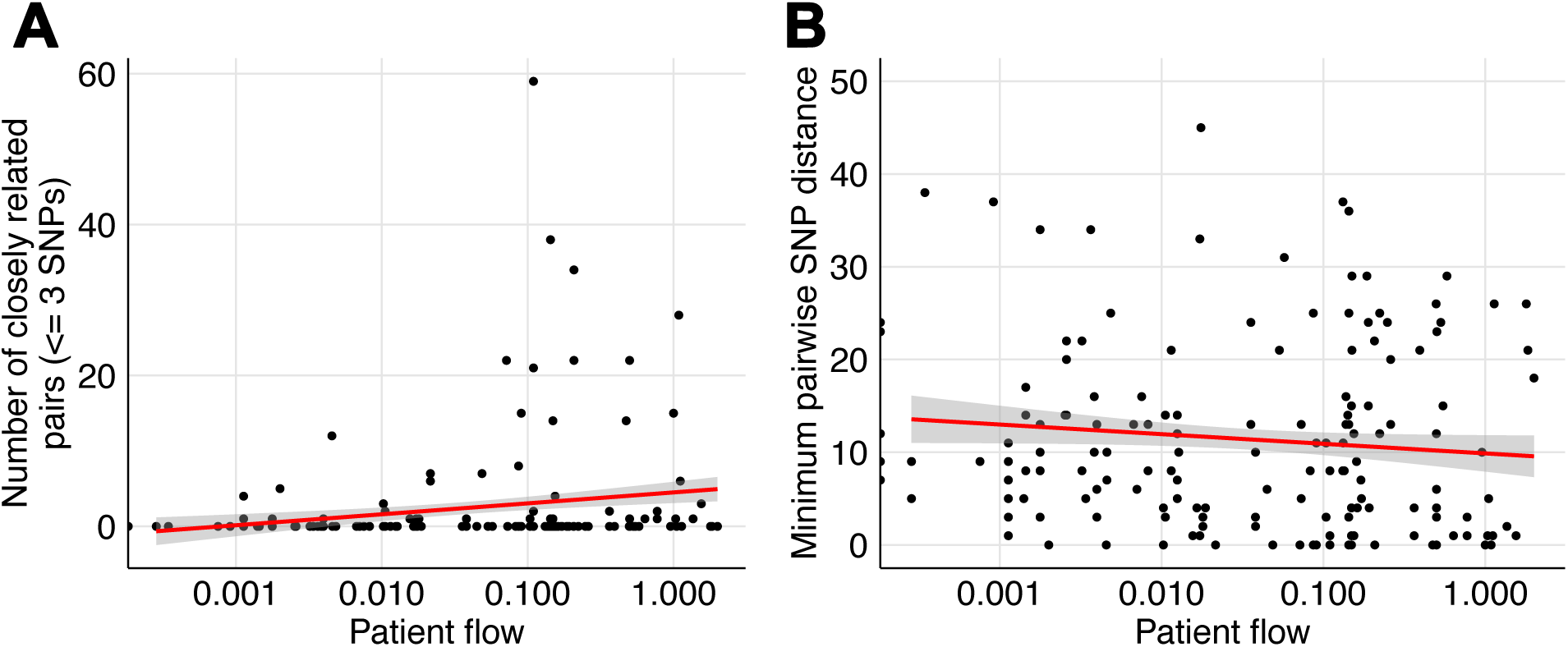
More frequent patient transfers are associated with more closely related *C. auris* genomes. Association between bi-directional patient flow, calculated from facility transfer networks, and (**A**) the number of inter-facility isolate pairs with 3 or few SNP differences, selected based on the median within cluster SNP distance or (**B**) the minimum pairwise SNP distance between inter-facility pairs.

### Analysis of isolates collected sequentially from individuals

Genomes of isolates collected from the same individual were usually closely related, with a median SNP difference of 5 (mean 8.3) compared to overall median SNP difference of 25 (mean 25.8) for all clade IV isolates (Fig. 2c). Forty-nine of 65 (75%) individuals were colonized with isolates over time that fell into single tree clades and/or had ≤7 SNP differences (Fig. 6a).

**Fig. 6.**
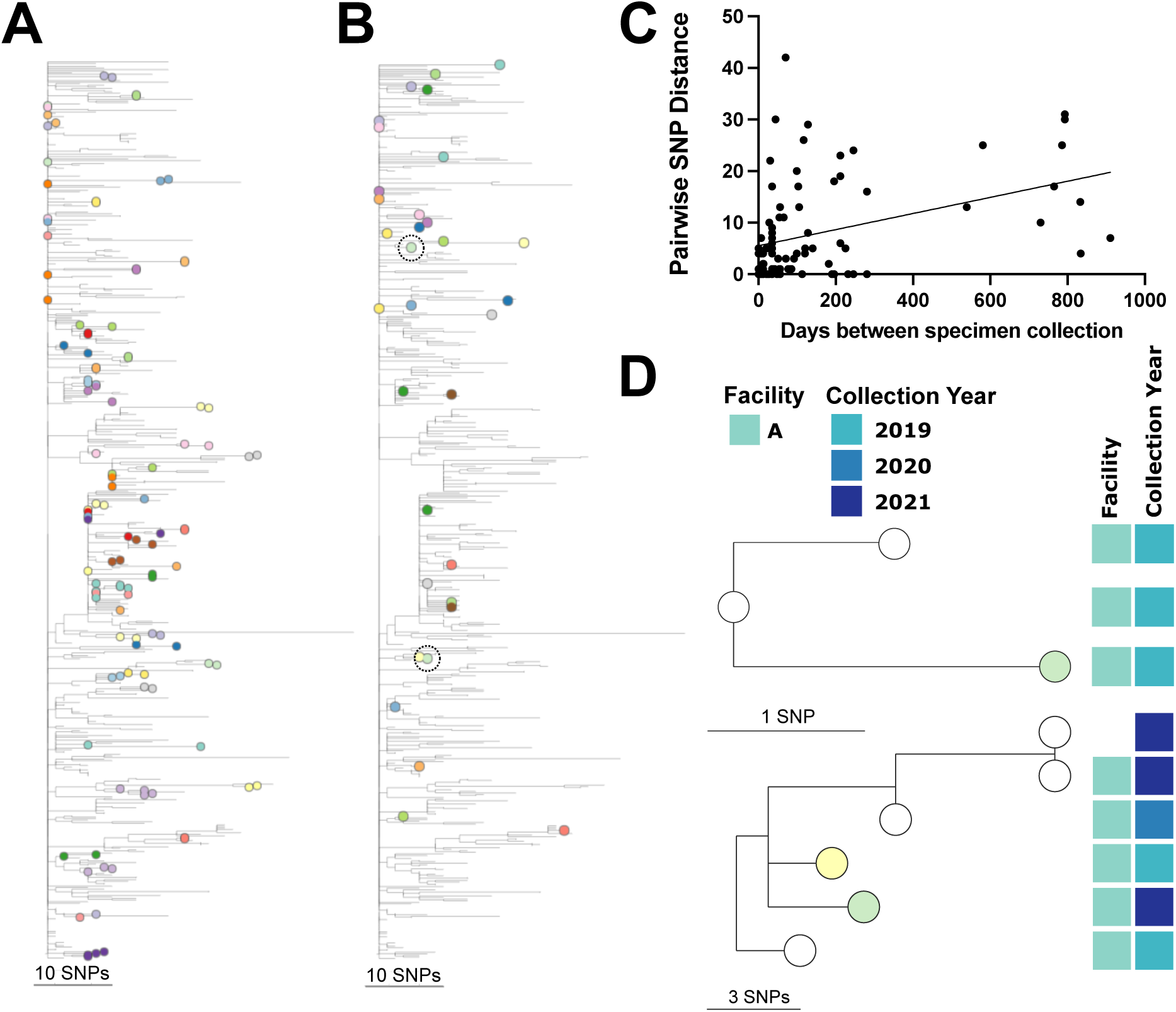
Relatedness of isolates collected from the same individual. (**A-B**) Phylogenetic tree as in Fig. 2a with tree tips colored by (**A**) individuals with isolates showing phylogenetic linkage and/or ≤ 7 SNP differences and (**B**) individuals with isolates showing divergent clustering and > 7 SNP differences. Isolates from one individual highlighted in subtrees in panel D are circled. (**C**) Correlation between pairwise SNP distance and the number of days between pairwise isolates collected longitudinally from one individual with linear regression. (**D**) Subtrees of one individual with divergent longitudinal isolates (circled in panel B) with relevant metadata in blocks to the right of tree tips. Tree tips indicate individuals as in panel B.

However, not all longitudinal isolates were closely related: pairwise SNP distances between isolates from the same individual ranged from 0-42 SNPs. Mean number of days between longitudinal isolate collections was 163 (range: 0-911 days); time between isolate collection and pairwise SNP distance was significantly correlated (Fig. 6b, Spearman r=0.34, p=0.0012). Within longitudinal series of isolates from the same patients, 42 of 65 (65%) were collected from individuals in the same facility and 22 (34%) were collected from the same patients in different facilities, exemplifying how *C. auris* may transfer between facilities.

All isolates from 18 (28%) of 65 individuals who were sampled repeatedly fell into a single genomic cluster, and 17 genomic clusters included only isolates from one individual. Another 17 (26%) individuals carried isolates longitudinally that fell together into a tree clade with ≤7 SNP differences. However, clustering with other more diverged isolates resulted in maximum genetic distances within the phylogenetic clade above our cutoff of 7 SNPs. Six (9.2%) individuals carried closely related isolates with ≤7 SNP differences over periods of 13 – 246 days but these isolates did not show tree linkage as they were only minimally diverged from the Illinois ancestor. Longitudinal isolates from 8 (12%) individuals showed shared branches but >7 SNP differences, indicating greater than expected genetic divergence. This could be due to long times between sampling resulting in higher divergence (2 of these 8 individuals were sampled >1 year apart) or falling into the same phylogenetic clade by chance. Sixteen (25%) longitudinally sampled individuals carried diverged isolates (11-42 SNPs) with distinct phylogenetic placement (Fig. 6c). Of these, 5 individuals carried isolates that showed phylogenetic linkage to other isolates collected from the same facility, suggesting colonization with multiple lineages circulating in the facility (Fig. 6d). In almost every case, closely related isolates from the same facility were collected within a few months of one another, suggesting colonization with contemporaneous isolates from the facility.

## DISCUSSION

Through integration of isolate WGS, patient transfer data, and epidemiologic metadata, we delineated the emergence and dissemination of *C. auris* across metropolitan Chicago beginning with its first detection in 2016 and continuing over five years. To our knowledge, this work is the first to incorporate bioinformatic cluster selection to identify closely related *C. auris* isolates and putative transmission clusters in large genomic datasets. Incorporating isolate and facility metadata allowed us to confirm linkage of isolates grouped into genomic clusters, identify facilities with evidence of intra- and inter-facility transmission, and estimate the relative contributions of intra- and inter-facility transmission to *C. auris* spread in Chicago, distinctions that are essential for designing effective mitigation strategies. Further, patient transfer data corroborated transmission linkages identified in genomic data, suggesting that it could be a useful tool for assessing inter-facility *C. auris* transmission risk. While others have described the genomic epidemiology of *C. auris* in outbreaks at the institutional and national scale, the current work shows that genomic analysis can inform understanding and response to *C. auris* spread at a regional level, informing prioritization of *C. auris* screening efforts (e.g., PPS and admission screening) and infection control recommendations.

Phylogenetic analysis of *C. auris* isolates identified a single introduction of clade IV to Illinois with local diversification and persistence of many lineages over six years. *C. auris* genotypes were intermixed across time and through many healthcare facilities. Like others, we found that healthcare facilities with *C. auris* are highly interconnected through patient transfer, likely contributing to dissemination of *C. auris* and strain sharing across facilities (*27*). WGS also identified a more recent introduction of clade III with brief local transmission, but no evidence for its continued dissemination.

WGS identified multiple isolate genome clusters with very close genetic relatedness that when supplemented with epidemiologic metadata provided evidence for both intra-facility and inter-facility transmission. Likely intra-facility transmission pairs were enriched in closely related isolates and found in 68% of facilities with sequenced *C. auris* isolates, but inter-facility transmission was also frequently found and contributed to 38% of closely related isolates. While closely related isolates were frequently found in genomic data, these likely transmission clusters were mainly limited to small groups of fewer than five individuals rather than larger outbreaks. Understanding transmission pathways of *C. auris* through integration of WGS and epidemiologic metadata can guide targeted interventions. For instance, proper hand hygiene and environmental cleaning with a disinfectant active against *C. auris* could mitigate spread within or between wards. In contrast, admission screening can be an effective method for facilities at risk for inter-facility transmission to rapidly identify and place colonized patients on the appropriate transmission-based precautions. Patient transfer data also corroborated transmission pathways identified through the genomic analysis in two of three large genomic clusters and facilities with more patient transfers had more closely related *C. auris* genome pairs. Thus, patient transfer data may serve as a predictive tool for inter-facility transmission of *C. auris,* allowing strategic prioritization of prevention efforts.

Bioinformatic classification of *C. auris* genomic clusters was employed here to help guide identification and description of related isolates in a large genomic dataset. There is no established SNP threshold for *C. auris* genomic/transmission linkage. Therefore, we set a reasonable SNP cut-off based on our own data, strengthened by the integration of relevant patient metadata. First, the SNP distance where enrichment with intra-facility isolate pairs declined and where transmission linkage is less likely was identified, as previously employed for bacterial SNP threshold determination (*26*). Second, we identified the SNP distance where the proportion of isolates collected from the same individual declined. This method is more conservative than SNP thresholds used in some bacterial studies, such as using maximal SNP distances within 95% of isolates from the same individual, which may not be useful for *C. auris* given that colonization with divergent genomes is seen in a large proportion of *C. auris* cases (*8, 28, 29*). Using either method, the SNP cut-off was ∼7 SNPs, which also corresponds approximately to SNPs expected to accumulate in a *C. auris* genome over 1 year (*8*). Furthermore, we employed phylogenetic clustering and maximum genetic divergence, rather than exclusively SNP-based cluster detection, given the relatively recent emergence of *C. auris* in Chicago and limited diversity observed in regional *C. auris* sequences, especially in the few years following local emergence. Thus, only genomic clusters with shared mutations diverging from the base of the Illinois phylogenetic clade were selected. Together, these genomic cluster thresholds aimed to identify close genetic relationships and recent transmission events. However, these conservative thresholds may have precluded identification of larger genomic clusters. For example, closely related isolates collected from the same individual often failed to be grouped by this cluster classification method when they fell into phylogenetic clades with other more diverse isolates that resulted in the clade exceeding the maximum genetic diversity threshold. Thus, minimum or mean genetic distance within monophyletic clades may be more useful for identifying *C. auris* genomic clusters involving many individuals or long-term ongoing transmission. Finally, myriad factors such as bioinformatic SNP-selection protocol, *C. auris* clade (I-VI), local transmission dynamics, and time since local *C. auris* emergence could affect appropriate SNP thresholds; these factors should be considered carefully when selecting SNP cutoffs for *C. auris* genomic analysis.

There were several limitations to this investigation. Most sequenced isolates were from four Chicago vSNFs or four Chicago LTACHs since CDPH conducts periodic PPSs in these facilities. Although a large proportion of Chicago *C. auris* cases are identified in acute care settings, associated isolates are often diagnosed internally and not sent routinely to a public health laboratory and therefore were not available for sequencing. Few isolates were available in 2020 as CDPH redirected resources from *C. auris* control to COVID-19 pandemic response.

Additionally, per laboratory protocols, isolates from Chicago PPSs are held at the WSLH for two years before release back to CDPH. At the time of the current analysis, isolates through 2021 were available for WGS. Finally, in most cases, only one isolate was available from each individual, limiting analyses of serial sampling. Others have shown that patients can be colonized with diverse genotypes of *C. auris* (*8, 29*). Indeed, we observed divergent *C. auris* genomes in 25% of individuals with multiple isolates collected, although available isolates were largely from serial and not single timepoints. This suggests that the *C. auris* isolates with which patients are colonized may be diverse or may change over time, especially in facilities in which *C. auris* is endemic. Thus, our approach of sequencing a single isolate from most individuals may have resulted in some genetic linkages being missed. Patient transfer data also included limitations, such as not providing a reason for facility transfer or a description of the type of transfer, which may impact the relevance of the number of transfers for *C. auris* transmission risk (e.g., patients with serious medical conditions or requiring a higher level of care may be most relevant for *C. auris*). Further, patient transfer data were not available for all included years; a single year’s data at the median collection date for included isolates was selected. Thus, some facilities connected by rare patient transfers may have been missed. We confirmed that facility transfer trends were consistent across years in available datasets. Finally, patient transfer data are derived from the Medicare and Medicaid population and are not representative of individuals in all payer networks.

In sum, this investigation utilized WGS, patient metadata, and patient transfer data to characterize the emergence of *C. auris* after a single introduction through dissemination and endemicity of *C. auris* in a new metropolitan area across 5 years. This work highlights the value of comprehensive *C. auris* genomic surveillance in public health programs and exemplifies the value of integrating multiple data types to characterize the rapidly evolving landscape of *C. auris* transmission. While *C. auris* WGS is often reserved for outbreak investigation, the actionable data informed by this retrospective genomic analysis supports timely and less focused use of WGS to support regional *C. auris* response. For example, untargeted WGS can enhance case-based surveillance by identifying genetically related isolates across facilities, thus identifying facilities at risk for inter-facility transmission for enhanced *C. auris* mitigation efforts, e.g. admission screening. We also investigated multiple data sources for their utility in informing *C. auris* transmission patterns; given that patient transfer information was concordant with WGS analysis, it should be considered as a predictive tool for risk of inter-facility transmission assessment.

## Data availability

All new sequencing data is available in the NCBI short read archive (SRA) under BioProject PRJNA904373, individual isolate accessions are listed in Supplemental Table S1. All external sequence data was downloaded from the NCBI SRA, accessions for all samples included in the analysis are listed in Supplemental Table S1. An interactive version of the Fig. 2 and Fig. 6 phylogeny is available at https://microreact.org/project/gV9gnmBgz18n8fjvyjxZtG.

## Conflicts of interest

Authors declare that they have no competing interests.

## Funding

This project was supported by the Centers for Disease Control and Prevention of the U.S. Department of Health and Human Services (HHS) as part of a financial assistance award totaling $11,162,000 with 100% funded by CDC/HHS. The contents are those of the author(s) and do not necessarily represent the official views of, nor an endorsement, by CDC/HHS, or the U.S. Government. The project described was supported in part by cooperative agreement U54 CK000607 from CDC. Its contents are solely the responsibility of the authors and do not necessarily represent the official views of CDC.

## Ethical approval and consent to participate

Informed consent for specimen collection was waived because collection was part of a public health response by the Chicago Department of Public Health under regulation 45 CFR 46.102(I)(2).

## Author contributions

Conceptualization: HJB, KAW, DYK, SRB, MKH

Methodology: HJB, KAW, EN, SJG

Investigation: HJB, KAW

Visualization: HJB, KAW

Funding acquisition: AK, SRB, SJG, MKH

Project administration: AK, CZ, HA

Supervision: HJB, AK, SJG, SRB, MKH

Writing – original draft: HJB, KAW

Writing – review & editing: DYK, SRB, MKH

## Acknowledgements

We thank members of the Wisconsin State Laboratory of Hygiene for coordination of isolates for sequencing. We thank members of the Rush University Medical Center Genomics and Microbiome Core Facility and Regional Innovative Public Health Laboratory for WGS, isolate and data management. We gratefully acknowledge the Chicago Department of Public Health Laboratory-Based Surveillance and Healthcare Settings teams.

## List of Supplementary Materials

Fig. S1 Table S1

## Notes

### Competing Interest Statement

The authors have declared no competing interest.

### Author Declarations

The study only used remnant already collected isolates. All samples were de-identified prior to inclusion. The Patient IDs were not disclosed to anyone outside of the research group.

