## Supplementary Information for "Transmission patterns of *Candida auris* among regional healthcare facilities revealed by genomic epidemiology and patient transfer data"

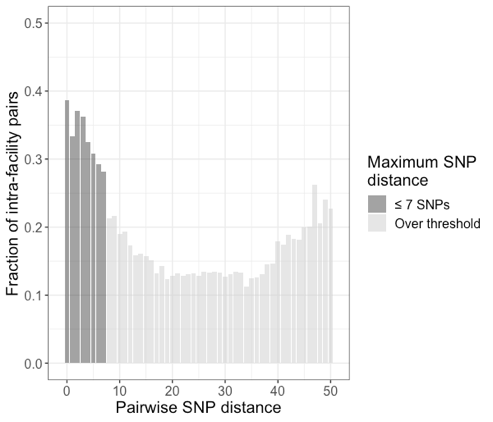

**Fig. S1. Maximum SNP distance selection based on SNP distance of intra-facility pairs.** The fraction of pairwise comparisons that are intra-facility at each pairwise SNP distance. Selected maximum SNP distance for cluster detection is shaded.

**Table S1. *Candida auris* sequence data included in genomic analysis**

| Sequence ID | SRA Accession | Sequence Type | C. auris clade |
| --- | --- | --- | --- |
| B11205 | SRR3883436 | Clade reference | I |
| B11220 | SRR3883452 | Clade reference | II |
| B11221 | SRR3883453 | Clade reference | III |
| B11243 | SRR3883464 | Clade reference | IV |
| B11842 | SRR7909141 | Illinois contextual | IV |
| B11843 | SRR7909220 | Illinois contextual | IV |
| B11880 | SRR7909238 | Illinois contextual | IV |
| B11882 | SRR7909309 | Illinois contextual | IV |
| B11883 | SRR7909391 | Illinois contextual | IV |
| B11884 | SRR7909250 | Illinois contextual | IV |
| B11885 | SRR7909191 | Illinois contextual | IV |
| B11886 | SRR7909407 | Illinois contextual | IV |
| B12028 | SRR7909256 | Illinois contextual | IV |
| B12029 | SRR7909248 | Illinois contextual | IV |
| B12030 | SRR7909228 | Illinois contextual | IV |
| B12031 | SRR7909172 | Illinois contextual | IV |
| B12032 | SRR7909387 | Illinois contextual | IV |
| B12033 | SRR7909179 | Illinois contextual | IV |
| B12034 | SRR7909388 | Illinois contextual | IV |
| B12035 | SRR7909335 | Illinois contextual | IV |
| B12036 | SRR7909151 | Illinois contextual | IV |
| B12046 | SRR7909366 | Illinois contextual | IV |
| B12077 | SRR7909138 | Illinois contextual | IV |
| B12189 | SRR7909158 | Illinois contextual | IV |
| B12190 | SRR7909245 | Illinois contextual | IV |
| B12229 | SRR7909200 | Illinois contextual | IV |
| B12378 | SRR7909145 | Illinois contextual | IV |
| B12380 | SRR7909294 | Illinois contextual | IV |
| B12388 | SRR7909221 | Illinois contextual | IV |
| B12406 | SRR7909405 | Illinois contextual | IV |
| B12510 | SRR7909227 | Illinois contextual | IV |
| B12579 | SRR7909315 | Illinois contextual | IV |
| B12937 | SRR7909226 | Illinois contextual | IV |
| B12938 | SRR7909357 | Illinois contextual | IV |
| B20665 | SRR23919790 | Illinois contextual | IV |
| B20666 | SRR23919789 | Illinois contextual | IV |
| B21737 | SRR23919786 | Illinois contextual | IV |
| B21738 | SRR23919785 | Illinois contextual | IV |
| B21739 | SRR23919784 | Illinois contextual | IV |
| B21740 | SRR23919783 | Illinois contextual | IV |
| B21741 | SRR23919782 | Illinois contextual | IV |
| B21742 | SRR23919781 | Illinois contextual | IV |
| B21743 | SRR23919780 | Illinois contextual | IV |
| B21744 | SRR23919779 | Illinois contextual | IV |
| B21745 | SRR23919788 | Illinois contextual | IV |
| B21746 | SRR23919787 | Illinois contextual | IV |
| CA01 | SRR12363130 | Illinois contextual | IV |
| CA02 | SRR12363129 | Illinois contextual | IV |
| CA03 | SRR12363163 | Illinois contextual | IV |
| CA04 | SRR12363162 | Illinois contextual | IV |
| CA05 | SRR12363161 | Illinois contextual | IV |
| CA06 | SRR12363160 | Illinois contextual | IV |
| CA07 | SRR12363159 | Illinois contextual | IV |
| CA08 | SRR12363158 | Illinois contextual | IV |
| CA09 | SRR12363157 | Illinois contextual | IV |
| CA10 | SRR12363156 | Illinois contextual | IV |
| CA11 | SRR12363155 | Illinois contextual | IV |
| CA12 | SRR12363154 | Illinois contextual | IV |
| CA13 | SRR12363152 | Illinois contextual | IV |
| CA14 | SRR12363151 | Illinois contextual | IV |
| CA15 | SRR12363150 | Illinois contextual | IV |
| CA16 | SRR12363149 | Illinois contextual | IV |
| CA17 | SRR12363148 | Illinois contextual | IV |
| CA18 | SRR12363147 | Illinois contextual | IV |
| CA19 | SRR12363146 | Illinois contextual | IV |
| CA20 | SRR12363145 | Illinois contextual | IV |
| CA21 | SRR12363144 | Illinois contextual | IV |
| CA22 | SRR12363143 | Illinois contextual | IV |
| CA23 | SRR12363141 | Illinois contextual | IV |
| CA24 | SRR12363140 | Illinois contextual | IV |
| CA25 | SRR12363139 | Illinois contextual | IV |
| IL-RIPHL_CAU-017070-0401 | SRR29662561 | New sequence | IV |
| IL-RIPHL_CAU-018070-0404 | SRR29662496 | New sequence | IV |
| IL-RIPHL_CAU-018070-0405 | SRR29662485 | New sequence | IV |
| IL-RIPHL_CAU-018070-0406 | SRR29662474 | New sequence | IV |
| IL-RIPHL_CAU-018070-0407 | SRR29662463 | New sequence | IV |
| IL-RIPHL_CAU-018070-0408 | SRR29662452 | New sequence | IV |
| IL-RIPHL_CAU-018070-0409 | SRR29662441 | New sequence | IV |
| IL-RIPHL_CAU-018070-0450 | SRR29662384 | New sequence | IV |
| IL-RIPHL_CAU-018070-0462 | SRR29662372 | New sequence | IV |
| IL-RIPHL_CAU-030070-0402 | SRR29662550 | New sequence | IV |
| IL-RIPHL_CAU-030070-0457 | SRR29662377 | New sequence | IV |
| IL-RIPHL_CAU-030070-0458 | SRR29662376 | New sequence | IV |
| IL-RIPHL_CAU-030070-0459 | SRR29662375 | New sequence | IV |
| IL-RIPHL_CAU-033070-0391 | SRR29662573 | New sequence | IV |
| IL-RIPHL_CAU-033070-0392 | SRR29662430 | New sequence | IV |
| IL-RIPHL_CAU-033070-0403 | SRR29662507 | New sequence | IV |
| IL-RIPHL_CAU-033070-0502 | SRR29662562 | New sequence | IV |
| IL-RIPHL_CAU-033070-0503 | SRR29662560 | New sequence | IV |
| IL-RIPHL_CAU-033070-0504 | SRR29662559 | New sequence | IV |
| IL-RIPHL_CAU-033070-0505 | SRR29662558 | New sequence | IV |
| IL-RIPHL_CAU-033070-0506 | SRR29662557 | New sequence | IV |
| IL-RIPHL_CAU-033070-0507 | SRR29662556 | New sequence | IV |
| IL-RIPHL_CAU-033070-0508 | SRR29662555 | New sequence | IV |
| IL-RIPHL_CAU-033070-0509 | SRR29662554 | New sequence | IV |
| IL-RIPHL_CAU-033070-0510 | SRR29662553 | New sequence | IV |
| IL-RIPHL_CAU-033070-0511 | SRR29662552 | New sequence | IV |
| IL-RIPHL_CAU-034070-0559 | SRR29662469 | New sequence | IV |
| IL-RIPHL_CAU-035070-0390 | SRR29662574 | New sequence | IV |
| IL-RIPHL_CAU-035070-0537 | SRR29662491 | New sequence | IV |
| IL-RIPHL_CAU-035070-0538 | SRR29662490 | New sequence | IV |
| IL-RIPHL_CAU-035070-0539 | SRR29662489 | New sequence | IV |
| IL-RIPHL_CAU-035070-0540 | SRR29662488 | New sequence | IV |
| IL-RIPHL_CAU-035070-0541 | SRR29662487 | New sequence | IV |
| IL-RIPHL_CAU-035070-0542 | SRR29662486 | New sequence | IV |
| IL-RIPHL_CAU-035070-0543 | SRR29662484 | New sequence | IV |
| IL-RIPHL_CAU-035070-0544 | SRR29662483 | New sequence | IV |
| IL-RIPHL_CAU-035070-0545 | SRR29662482 | New sequence | IV |
| IL-RIPHL_CAU-035070-0546 | SRR29662481 | New sequence | IV |
| IL-RIPHL_CAU-035070-0547 | SRR29662480 | New sequence | IV |
| IL-RIPHL_CAU-035070-0548 | SRR29662479 | New sequence | IV |
| IL-RIPHL_CAU-035070-0549 | SRR29662478 | New sequence | IV |
| IL-RIPHL_CAU-035070-0550 | SRR29662477 | New sequence | IV |
| IL-RIPHL_CAU-035070-0551 | SRR29662476 | New sequence | IV |
| IL-RIPHL_CAU-035070-0552 | SRR29662475 | New sequence | IV |
| IL-RIPHL_CAU-035070-0553 | SRR29662473 | New sequence | IV |
| IL-RIPHL_CAU-035070-0554 | SRR29662472 | New sequence | IV |
| IL-RIPHL_CAU-035070-0555 | SRR29662471 | New sequence | IV |
| IL-RIPHL_CAU-036070-0529 | SRR29662500 | New sequence | IV |
| IL-RIPHL_CAU-036070-0530 | SRR29662499 | New sequence | IV |
| IL-RIPHL_CAU-036070-0531 | SRR29662498 | New sequence | IV |
| IL-RIPHL_CAU-036070-0532 | SRR29662497 | New sequence | IV |
| IL-RIPHL_CAU-036070-0533 | SRR29662495 | New sequence | IV |
| IL-RIPHL_CAU-036070-0534 | SRR29662494 | New sequence | IV |
| IL-RIPHL_CAU-036070-0535 | SRR29662493 | New sequence | IV |
| IL-RIPHL_CAU-036070-0536 | SRR29662492 | New sequence | IV |
| IL-RIPHL_CAU-037070-0397 | SRR29662371 | New sequence | IV |
| IL-RIPHL_CAU-037070-0398 | SRR29662360 | New sequence | IV |
| IL-RIPHL_CAU-037070-0399 | SRR29662349 | New sequence | IV |
| IL-RIPHL_CAU-037070-0400 | SRR29662572 | New sequence | IV |
| IL-RIPHL_CAU-037070-0410 | SRR29662429 | New sequence | IV |
| IL-RIPHL_CAU-037070-0412 | SRR29662418 | New sequence | IV |
| IL-RIPHL_CAU-037070-0414 | SRR29662535 | New sequence | IV |
| IL-RIPHL_CAU-037070-0415 | SRR29662524 | New sequence | IV |
| IL-RIPHL_CAU-037070-0416 | SRR29662517 | New sequence | IV |
| IL-RIPHL_CAU-037070-0417 | SRR29662516 | New sequence | IV |
| IL-RIPHL_CAU-037070-0418 | SRR29662515 | New sequence | IV |
| IL-RIPHL_CAU-037070-0419 | SRR29662514 | New sequence | IV |
| IL-RIPHL_CAU-037070-0420 | SRR29662513 | New sequence | IV |
| IL-RIPHL_CAU-037070-0421 | SRR29662512 | New sequence | IV |
| IL-RIPHL_CAU-037070-0422 | SRR29662414 | New sequence | IV |
| IL-RIPHL_CAU-037070-0423 | SRR29662413 | New sequence | IV |
| IL-RIPHL_CAU-037070-0424 | SRR29662412 | New sequence | IV |
| IL-RIPHL_CAU-037070-0425 | SRR29662411 | New sequence | IV |
| IL-RIPHL_CAU-037070-0426 | SRR29662410 | New sequence | IV |
| IL-RIPHL_CAU-037070-0427 | SRR29662409 | New sequence | IV |
| IL-RIPHL_CAU-037070-0428 | SRR29662408 | New sequence | IV |
| IL-RIPHL_CAU-037070-0429 | SRR29662407 | New sequence | IV |
| IL-RIPHL_CAU-037070-0430 | SRR29662406 | New sequence | IV |
| IL-RIPHL_CAU-037070-0431 | SRR29662405 | New sequence | IV |
| IL-RIPHL_CAU-037070-0432 | SRR29662403 | New sequence | IV |
| IL-RIPHL_CAU-037070-0433 | SRR29662402 | New sequence | IV |
| IL-RIPHL_CAU-037070-0434 | SRR29662401 | New sequence | IV |
| IL-RIPHL_CAU-037070-0435 | SRR29662400 | New sequence | IV |
| IL-RIPHL_CAU-037070-0436 | SRR29662399 | New sequence | IV |
| IL-RIPHL_CAU-037070-0437 | SRR29662398 | New sequence | IV |
| IL-RIPHL_CAU-037070-0438 | SRR29662397 | New sequence | IV |
| IL-RIPHL_CAU-037070-0439 | SRR29662396 | New sequence | IV |
| IL-RIPHL_CAU-037070-0440 | SRR29662395 | New sequence | IV |
| IL-RIPHL_CAU-037070-0441 | SRR29662394 | New sequence | IV |
| IL-RIPHL_CAU-037070-0442 | SRR29662392 | New sequence | IV |
| IL-RIPHL_CAU-037070-0443 | SRR29662391 | New sequence | IV |
| IL-RIPHL_CAU-037070-0444 | SRR29662390 | New sequence | IV |
| IL-RIPHL_CAU-037070-0445 | SRR29662389 | New sequence | IV |
| IL-RIPHL_CAU-037070-0446 | SRR29662388 | New sequence | IV |
| IL-RIPHL_CAU-037070-0447 | SRR29662387 | New sequence | IV |
| IL-RIPHL_CAU-037070-0448 | SRR29662386 | New sequence | IV |
| IL-RIPHL_CAU-037070-0449 | SRR29662385 | New sequence | IV |
| IL-RIPHL_CAU-037070-0451 | SRR29662383 | New sequence | IV |
| IL-RIPHL_CAU-037070-0453 | SRR29662381 | New sequence | IV |
| IL-RIPHL_CAU-037070-0454 | SRR29662380 | New sequence | IV |
| IL-RIPHL_CAU-037070-0455 | SRR29662379 | New sequence | IV |
| IL-RIPHL_CAU-037070-0456 | SRR29662378 | New sequence | IV |
| IL-RIPHL_CAU-037070-0460 | SRR29662374 | New sequence | IV |
| IL-RIPHL_CAU-037070-0461 | SRR29662373 | New sequence | IV |
| IL-RIPHL_CAU-037070-0463 | SRR29662370 | New sequence | IV |
| IL-RIPHL_CAU-037070-0464 | SRR29662369 | New sequence | IV |
| IL-RIPHL_CAU-037070-0465 | SRR29662368 | New sequence | IV |
| IL-RIPHL_CAU-037070-0466 | SRR29662367 | New sequence | IV |
| IL-RIPHL_CAU-037070-0467 | SRR29662366 | New sequence | IV |
| IL-RIPHL_CAU-037070-0468 | SRR29662365 | New sequence | IV |
| IL-RIPHL_CAU-037070-0469 | SRR29662364 | New sequence | IV |
| IL-RIPHL_CAU-037070-0470 | SRR29662363 | New sequence | IV |
| IL-RIPHL_CAU-037070-0471 | SRR29662362 | New sequence | IV |
| IL-RIPHL_CAU-037070-0472 | SRR29662361 | New sequence | IV |
| IL-RIPHL_CAU-037070-0473 | SRR29662359 | New sequence | IV |
| IL-RIPHL_CAU-037070-0474 | SRR29662358 | New sequence | IV |
| IL-RIPHL_CAU-037070-0475 | SRR29662357 | New sequence | IV |
| IL-RIPHL_CAU-037070-0476 | SRR29662356 | New sequence | IV |
| IL-RIPHL_CAU-037070-0477 | SRR29662355 | New sequence | IV |
| IL-RIPHL_CAU-037070-0478 | SRR29662354 | New sequence | IV |
| IL-RIPHL_CAU-037070-0479 | SRR29662353 | New sequence | IV |
| IL-RIPHL_CAU-037070-0480 | SRR29662352 | New sequence | IV |
| IL-RIPHL_CAU-037070-0481 | SRR29662351 | New sequence | IV |
| IL-RIPHL_CAU-037070-0482 | SRR29662350 | New sequence | IV |
| IL-RIPHL_CAU-037070-0483 | SRR29662348 | New sequence | IV |
| IL-RIPHL_CAU-037070-0484 | SRR29662347 | New sequence | IV |
| IL-RIPHL_CAU-037070-0485 | SRR29662346 | New sequence | IV |
| IL-RIPHL_CAU-037070-0486 | SRR29662345 | New sequence | IV |
| IL-RIPHL_CAU-037070-0487 | SRR29662344 | New sequence | IV |
| IL-RIPHL_CAU-037070-0488 | SRR29662343 | New sequence | IV |
| IL-RIPHL_CAU-037070-0489 | SRR29662342 | New sequence | IV |
| IL-RIPHL_CAU-037070-0490 | SRR29662341 | New sequence | IV |
| IL-RIPHL_CAU-037070-0491 | SRR29662340 | New sequence | IV |
| IL-RIPHL_CAU-037070-0492 | SRR29662339 | New sequence | IV |
| IL-RIPHL_CAU-037070-0493 | SRR29662571 | New sequence | IV |
| IL-RIPHL_CAU-037070-0494 | SRR29662570 | New sequence | IV |
| IL-RIPHL_CAU-037070-0495 | SRR29662569 | New sequence | IV |
| IL-RIPHL_CAU-037070-0496 | SRR29662568 | New sequence | IV |
| IL-RIPHL_CAU-037070-0497 | SRR29662567 | New sequence | IV |
| IL-RIPHL_CAU-037070-0498 | SRR29662566 | New sequence | IV |
| IL-RIPHL_CAU-037070-0499 | SRR29662565 | New sequence | IV |
| IL-RIPHL_CAU-037070-0500 | SRR29662564 | New sequence | IV |
| IL-RIPHL_CAU-037070-0501 | SRR29662563 | New sequence | IV |
| IL-RIPHL_CAU-037070-0526 | SRR29662503 | New sequence | IV |
| IL-RIPHL_CAU-037070-0527 | SRR29662502 | New sequence | IV |
| IL-RIPHL_CAU-037070-0528 | SRR29662501 | New sequence | IV |
| IL-RIPHL_CAU-037070-0558 | SRR29662470 | New sequence | IV |
| IL-RIPHL_CAU-037070-0560 | SRR29662468 | New sequence | IV |
| IL-RIPHL_CAU-037070-0561 | SRR29662467 | New sequence | IV |
| IL-RIPHL_CAU-037070-0562 | SRR29662466 | New sequence | IV |
| IL-RIPHL_CAU-037070-0563 | SRR29662465 | New sequence | IV |
| IL-RIPHL_CAU-037070-0564 | SRR29662464 | New sequence | IV |
| IL-RIPHL_CAU-037070-0565 | SRR29662462 | New sequence | IV |
| IL-RIPHL_CAU-037070-0566 | SRR29662461 | New sequence | IV |
| IL-RIPHL_CAU-037070-0567 | SRR29662460 | New sequence | IV |
| IL-RIPHL_CAU-037070-0568 | SRR29662459 | New sequence | IV |
| IL-RIPHL_CAU-037070-0569 | SRR29662458 | New sequence | IV |
| IL-RIPHL_CAU-037070-0570 | SRR29662457 | New sequence | IV |
| IL-RIPHL_CAU-037070-0571 | SRR29662456 | New sequence | IV |
| IL-RIPHL_CAU-037070-0572 | SRR29662455 | New sequence | IV |
| IL-RIPHL_CAU-037070-0573 | SRR29662454 | New sequence | IV |
| IL-RIPHL_CAU-037070-0574 | SRR29662453 | New sequence | IV |
| IL-RIPHL_CAU-037070-0575 | SRR29662451 | New sequence | IV |
| IL-RIPHL_CAU-037070-0576 | SRR29662450 | New sequence | IV |
| IL-RIPHL_CAU-037070-0577 | SRR29662449 | New sequence | IV |
| IL-RIPHL_CAU-037070-0578 | SRR29662448 | New sequence | IV |
| IL-RIPHL_CAU-037070-0579 | SRR29662447 | New sequence | IV |
| IL-RIPHL_CAU-037070-0597 | SRR29662426 | New sequence | IV |
| IL-RIPHL_CAU-037070-0598 | SRR29662425 | New sequence | IV |
| IL-RIPHL_CAU-037070-0599 | SRR29662424 | New sequence | IV |
| IL-RIPHL_CAU-037070-0600 | SRR29662423 | New sequence | IV |
| IL-RIPHL_CAU-037070-0601 | SRR29662422 | New sequence | IV |
| IL-RIPHL_CAU-037070-0602 | SRR29662421 | New sequence | IV |
| IL-RIPHL_CAU-037070-0603 | SRR29662420 | New sequence | IV |
| IL-RIPHL_CAU-037070-0604 | SRR29662419 | New sequence | IV |
| IL-RIPHL_CAU-037070-0605 | SRR29662417 | New sequence | IV |
| IL-RIPHL_CAU-037070-0606 | SRR29662416 | New sequence | IV |
| IL-RIPHL_CAU-037070-0607 | SRR29662415 | New sequence | IV |
| IL-RIPHL_CAU-037070-0608 | SRR29662542 | New sequence | IV |
| IL-RIPHL_CAU-037070-0609 | SRR29662541 | New sequence | IV |
| IL-RIPHL_CAU-037070-0610 | SRR29662540 | New sequence | IV |
| IL-RIPHL_CAU-037070-0611 | SRR29662539 | New sequence | IV |
| IL-RIPHL_CAU-037070-0612 | SRR29662538 | New sequence | IV |
| IL-RIPHL_CAU-037070-0613 | SRR29662537 | New sequence | IV |
| IL-RIPHL_CAU-037070-0614 | SRR29662536 | New sequence | IV |
| IL-RIPHL_CAU-037070-0615 | SRR29662534 | New sequence | IV |
| IL-RIPHL_CAU-037070-0616 | SRR29662533 | New sequence | IV |
| IL-RIPHL_CAU-037070-0617 | SRR29662532 | New sequence | IV |
| IL-RIPHL_CAU-037070-0618 | SRR29662531 | New sequence | IV |
| IL-RIPHL_CAU-037070-0619 | SRR29662530 | New sequence | IV |
| IL-RIPHL_CAU-037070-0620 | SRR29662529 | New sequence | IV |
| IL-RIPHL_CAU-037070-0621 | SRR29662528 | New sequence | IV |
| IL-RIPHL_CAU-037070-0622 | SRR29662527 | New sequence | IV |
| IL-RIPHL_CAU-037070-0623 | SRR29662526 | New sequence | IV |
| IL-RIPHL_CAU-037070-0624 | SRR29662525 | New sequence | IV |
| IL-RIPHL_CAU-037070-0625 | SRR29662523 | New sequence | IV |
| IL-RIPHL_CAU-038070-0580 | SRR29662446 | New sequence | IV |
| IL-RIPHL_CAU-038070-0581 | SRR29662445 | New sequence | IV |
| IL-RIPHL_CAU-038070-0582 | SRR29662444 | New sequence | IV |
| IL-RIPHL_CAU-038070-0584 | SRR29662442 | New sequence | IV |
| IL-RIPHL_CAU-038070-0585 | SRR29662440 | New sequence | IV |
| IL-RIPHL_CAU-038070-0586 | SRR29662439 | New sequence | IV |
| IL-RIPHL_CAU-038070-0587 | SRR29662438 | New sequence | IV |
| IL-RIPHL_CAU-038070-0588 | SRR29662437 | New sequence | IV |
| IL-RIPHL_CAU-038070-0589 | SRR29662436 | New sequence | IV |
| IL-RIPHL_CAU-038070-0590 | SRR29662435 | New sequence | IV |
| IL-RIPHL_CAU-038070-0592 | SRR29662433 | New sequence | IV |
| IL-RIPHL_CAU-038070-0594 | SRR29662431 | New sequence | IV |
| IL-RIPHL_CAU-038070-0595 | SRR29662428 | New sequence | IV |
| IL-RIPHL_CAU-038070-0596 | SRR29662427 | New sequence | IV |
| IL-RIPHL_CAU-039070-0393 | SRR29662511 | New sequence | IV |
| IL-RIPHL_CAU-039070-0394 | SRR29662404 | New sequence | IV |
| IL-RIPHL_CAU-039070-0395 | SRR29662393 | New sequence | IV |
| IL-RIPHL_CAU-039070-0396 | SRR29662382 | New sequence | IV |
| IL-RIPHL_CAU-039070-0512 | SRR29662551 | New sequence | IV |
| IL-RIPHL_CAU-039070-0513 | SRR29662549 | New sequence | IV |
| IL-RIPHL_CAU-039070-0514 | SRR29662548 | New sequence | IV |
| IL-RIPHL_CAU-039070-0515 | SRR29662547 | New sequence | IV |
| IL-RIPHL_CAU-039070-0516 | SRR29662546 | New sequence | IV |
| IL-RIPHL_CAU-039070-0517 | SRR29662545 | New sequence | IV |
| IL-RIPHL_CAU-039070-0518 | SRR29662544 | New sequence | IV |
| IL-RIPHL_CAU-039070-0519 | SRR29662543 | New sequence | IV |
| IL-RIPHL_CAU-039070-0520 | SRR29662510 | New sequence | IV |
| IL-RIPHL_CAU-039070-0521 | SRR29662509 | New sequence | IV |
| IL-RIPHL_CAU-039070-0522 | SRR29662508 | New sequence | IV |
| IL-RIPHL_CAU-039070-0523 | SRR29662506 | New sequence | IV |
| IL-RIPHL_CAU-039070-0524 | SRR29662505 | New sequence | IV |
| IL-RIPHL_CAU-039070-0525 | SRR29662504 | New sequence | IV |
| IL-RIPHL-CAU-012012-0386 | N/A | Illinois contextual |  |
| IL-RIPHL-CAU-012012-0387 | N/A | Illinois contextual |  |
| IL-RIPHL-CAU-012012-0388 | N/A | Illinois contextual |  |
| IL-RIPHL-CAU-012012-0389 | N/A | Illinois contextual |  |
| IL-RIPHL-CAU-120002 | SRR22393499 | Illinois contextual |  |
| IL-RIPHL-CAU-120003 | SRR22393498 | Illinois contextual |  |
| IL-RIPHL-CAU-120013 | SRR24673390 | Illinois contextual |  |
| IL-RIPHL-CAU-120015 | SRR24673377 | Illinois contextual |  |
| IL-RIPHL-CAU-120019 | SRR24673432 | Illinois contextual |  |
| IL-RIPHL-CAU-120021 | SRR24673430 | Illinois contextual |  |
| IL-RIPHL-CAU-120025 | SRR24673426 | Illinois contextual |  |
| IL-RIPHL-CAU-120032 | SRR24673418 | Illinois contextual |  |
| IL-RIPHL-CAU-120033 | SRR24673417 | Illinois contextual |  |
| IL-RIPHL-CAU-120050 | SRR24673398 | Illinois contextual |  |
| IL-RIPHL-CAU-120062 | SRR24673386 | Illinois contextual |  |
| IL-RIPHL-CAU-70-0070 | SRR27223802 | New sequence | IV |
| IL-RIPHL-CAU-70-0071 | SRR27223801 | New sequence | IV |
| IL-RIPHL-CAU-70-0072 | SRR27223572 | New sequence | IV |
| IL-RIPHL-CAU-70-0073 | SRR27223611 | New sequence | IV |
| IL-RIPHL-CAU-70-0074 | SRR27223600 | New sequence | IV |
| IL-RIPHL-CAU-70-0075 | SRR27223589 | New sequence | IV |
| IL-RIPHL-CAU-70-0076 | SRR27223546 | New sequence | IV |
| IL-RIPHL-CAU-70-0077 | SRR27223535 | New sequence | IV |
| IL-RIPHL-CAU-70-0078 | SRR27223738 | New sequence | IV |
| IL-RIPHL-CAU-70-0079 | SRR27223727 | New sequence | IV |
| IL-RIPHL-CAU-70-0080 | SRR27223662 | New sequence | IV |
| IL-RIPHL-CAU-70-0081 | SRR27223800 | New sequence | IV |
| IL-RIPHL-CAU-70-0082 | SRR27223789 | New sequence | IV |
| IL-RIPHL-CAU-70-0083 | SRR27223778 | New sequence | IV |
| IL-RIPHL-CAU-70-0084 | SRR27223713 | New sequence | IV |
| IL-RIPHL-CAU-70-0085 | SRR27223702 | New sequence | IV |
| IL-RIPHL-CAU-70-0086 | SRR27223691 | New sequence | IV |
| IL-RIPHL-CAU-70-0087 | SRR27223648 | New sequence | IV |
| IL-RIPHL-CAU-70-0088 | SRR27223637 | New sequence | IV |
| IL-RIPHL-CAU-70-0089 | SRR27223626 | New sequence | IV |
| IL-RIPHL-CAU-70-0090 | SRR27223583 | New sequence | IV |
| IL-RIPHL-CAU-70-0091 | SRR27223571 | New sequence | IV |
| IL-RIPHL-CAU-70-0092 | SRR27223560 | New sequence | IV |
| IL-RIPHL-CAU-70-0093 | SRR27223763 | New sequence | IV |
| IL-RIPHL-CAU-70-0094 | SRR27223752 | New sequence | IV |
| IL-RIPHL-CAU-70-0095 | SRR27223741 | New sequence | IV |
| IL-RIPHL-CAU-70-0096 | SRR27223676 | New sequence | IV |
| IL-RIPHL-CAU-70-0097 | SRR27223665 | New sequence | IV |
| IL-RIPHL-CAU-70-0098 | SRR27223654 | New sequence | IV |
| IL-RIPHL-CAU-70-0099 | SRR27223661 | New sequence | IV |
| IL-RIPHL-CAU-70-0100 | SRR27223613 | New sequence | IV |
| IL-RIPHL-CAU-70-0101 | SRR27223612 | New sequence | IV |
| IL-RIPHL-CAU-70-0102 | SRR27223610 | New sequence | IV |
| IL-RIPHL-CAU-70-0103 | SRR27223609 | New sequence | IV |
| IL-RIPHL-CAU-70-0104 | SRR27223608 | New sequence | IV |
| IL-RIPHL-CAU-70-0105 | SRR27223607 | New sequence | IV |
| IL-RIPHL-CAU-70-0106 | SRR27223606 | New sequence | IV |
| IL-RIPHL-CAU-70-0107 | SRR27223605 | New sequence | IV |
| IL-RIPHL-CAU-70-0108 | SRR27223604 | New sequence | IV |
| IL-RIPHL-CAU-70-0109 | SRR27223603 | New sequence | IV |
| IL-RIPHL-CAU-70-0110 | SRR27223602 | New sequence | IV |
| IL-RIPHL-CAU-70-0111 | SRR27223601 | New sequence | IV |
| IL-RIPHL-CAU-70-0112 | SRR27223599 | New sequence | IV |
| IL-RIPHL-CAU-70-0113 | SRR27223598 | New sequence | IV |
| IL-RIPHL-CAU-70-0114 | SRR27223597 | New sequence | IV |
| IL-RIPHL-CAU-70-0115 | SRR27223596 | New sequence | IV |
| IL-RIPHL-CAU-70-0116 | SRR27223595 | New sequence | IV |
| IL-RIPHL-CAU-70-0117 | SRR27223594 | New sequence | IV |
| IL-RIPHL-CAU-70-0118 | SRR27223593 | New sequence | IV |
| IL-RIPHL-CAU-70-0119 | SRR27223592 | New sequence | IV |
| IL-RIPHL-CAU-70-0120 | SRR27223591 | New sequence | IV |
| IL-RIPHL-CAU-70-0121 | SRR27223590 | New sequence | IV |
| IL-RIPHL-CAU-70-0122 | SRR27223556 | New sequence | IV |
| IL-RIPHL-CAU-70-0123 | SRR27223555 | New sequence | IV |
| IL-RIPHL-CAU-70-0124 | SRR27223554 | New sequence | IV |
| IL-RIPHL-CAU-70-0125 | SRR27223553 | New sequence | IV |
| IL-RIPHL-CAU-70-0126 | SRR27223552 | New sequence | IV |
| IL-RIPHL-CAU-70-0127 | SRR27223551 | New sequence | IV |
| IL-RIPHL-CAU-70-0128 | SRR27223550 | New sequence | IV |
| IL-RIPHL-CAU-70-0129 | SRR27223549 | New sequence | IV |
| IL-RIPHL-CAU-70-0130 | SRR27223548 | New sequence | IV |
| IL-RIPHL-CAU-70-0131 | SRR27223547 | New sequence | IV |
| IL-RIPHL-CAU-70-0132 | SRR27223545 | New sequence | IV |
| IL-RIPHL-CAU-70-0134 | SRR27223544 | New sequence | IV |
| IL-RIPHL-CAU-70-0135 | SRR27223543 | New sequence | IV |
| IL-RIPHL-CAU-70-0136 | SRR27223542 | New sequence | IV |
| IL-RIPHL-CAU-70-0137 | SRR27223541 | New sequence | IV |
| IL-RIPHL-CAU-70-0138 | SRR27223540 | New sequence | IV |
| IL-RIPHL-CAU-70-0139 | SRR27223539 | New sequence | IV |
| IL-RIPHL-CAU-70-0140 | SRR27223538 | New sequence | IV |
| IL-RIPHL-CAU-70-0141 | SRR27223537 | New sequence | IV |
| IL-RIPHL-CAU-70-0142 | SRR27223536 | New sequence | IV |
| IL-RIPHL-CAU-70-0143 | SRR27223534 | New sequence | IV |
| IL-RIPHL-CAU-70-0144 | SRR27223533 | New sequence | IV |
| IL-RIPHL-CAU-70-0145 | SRR27223532 | New sequence | IV |
| IL-RIPHL-CAU-70-0146 | SRR27223531 | New sequence | IV |
| IL-RIPHL-CAU-70-0147 | SRR27223530 | New sequence | IV |
| IL-RIPHL-CAU-70-0148 | SRR27223529 | New sequence | IV |
| IL-RIPHL-CAU-70-0149 | SRR27223528 | New sequence | IV |
| IL-RIPHL-CAU-70-0150 | SRR27223527 | New sequence | IV |
| IL-RIPHL-CAU-70-0151 | SRR27223526 | New sequence | IV |
| IL-RIPHL-CAU-70-0152 | SRR27223525 | New sequence | IV |
| IL-RIPHL-CAU-70-0153 | SRR27223737 | New sequence | IV |
| IL-RIPHL-CAU-70-0154 | SRR27223736 | New sequence | IV |
| IL-RIPHL-CAU-70-0155 | SRR27223660 | New sequence | IV |
| IL-RIPHL-CAU-70-0156 | SRR27223735 | New sequence | IV |
| IL-RIPHL-CAU-70-0157 | SRR27223734 | New sequence | IV |
| IL-RIPHL-CAU-70-0158 | SRR27223733 | New sequence | IV |
| IL-RIPHL-CAU-70-0159 | SRR27223732 | New sequence | IV |
| IL-RIPHL-CAU-70-0160 | SRR27223731 | New sequence | IV |
| IL-RIPHL-CAU-70-0161 | SRR27223730 | New sequence | IV |
| IL-RIPHL-CAU-70-0162 | SRR27223729 | New sequence | IV |
| IL-RIPHL-CAU-70-0163 | SRR27223728 | New sequence | IV |
| IL-RIPHL-CAU-70-0164 | SRR27223726 | New sequence | IV |
| IL-RIPHL-CAU-70-0166 | SRR27223725 | New sequence | IV |
| IL-RIPHL-CAU-70-0167 | SRR27223724 | New sequence | IV |
| IL-RIPHL-CAU-70-0168 | SRR27223723 | New sequence | IV |
| IL-RIPHL-CAU-70-0169 | SRR27223722 | New sequence | IV |
| IL-RIPHL-CAU-70-0170 | SRR27223721 | New sequence | IV |
| IL-RIPHL-CAU-70-0171 | SRR27223720 | New sequence | IV |
| IL-RIPHL-CAU-70-0172 | SRR27223719 | New sequence | IV |
| IL-RIPHL-CAU-70-0173 | SRR27223718 | New sequence | IV |
| IL-RIPHL-CAU-70-0174 | SRR27223717 | New sequence | IV |
| IL-RIPHL-CAU-70-0175 | SRR27223799 | New sequence | IV |
| IL-RIPHL-CAU-70-0176 | SRR27223798 | New sequence | IV |
| IL-RIPHL-CAU-70-0177 | SRR27223797 | New sequence | IV |
| IL-RIPHL-CAU-70-0178 | SRR27223796 | New sequence | IV |
| IL-RIPHL-CAU-70-0179 | SRR27223795 | New sequence | IV |
| IL-RIPHL-CAU-70-0180 | SRR27223794 | New sequence | IV |
| IL-RIPHL-CAU-70-0181 | SRR27223793 | New sequence | IV |
| IL-RIPHL-CAU-70-0182 | SRR27223792 | New sequence | IV |
| IL-RIPHL-CAU-70-0183 | SRR27223791 | New sequence | IV |
| IL-RIPHL-CAU-70-0184 | SRR27223790 | New sequence | IV |
| IL-RIPHL-CAU-70-0185 | SRR27223788 | New sequence | IV |
| IL-RIPHL-CAU-70-0186 | SRR27223787 | New sequence | IV |
| IL-RIPHL-CAU-70-0187 | SRR27223786 | New sequence | IV |
| IL-RIPHL-CAU-70-0188 | SRR27223785 | New sequence | IV |
| IL-RIPHL-CAU-70-0189 | SRR27223784 | New sequence | IV |
| IL-RIPHL-CAU-70-0190 | SRR27223783 | New sequence | IV |
| IL-RIPHL-CAU-70-0191 | SRR27223782 | New sequence | IV |
| IL-RIPHL-CAU-70-0192 | SRR27223781 | New sequence | IV |
| IL-RIPHL-CAU-70-0193 | SRR27223780 | New sequence | IV |
| IL-RIPHL-CAU-70-0194 | SRR27223779 | New sequence | IV |
| IL-RIPHL-CAU-70-0195 | SRR27223777 | New sequence | IV |
| IL-RIPHL-CAU-70-0196 | SRR27223776 | New sequence | IV |
| IL-RIPHL-CAU-70-0197 | SRR27223775 | New sequence | IV |
| IL-RIPHL-CAU-70-0198 | SRR27223774 | New sequence | IV |
| IL-RIPHL-CAU-70-0199 | SRR27223773 | New sequence | IV |
| IL-RIPHL-CAU-70-0200 | SRR27223772 | New sequence | IV |
| IL-RIPHL-CAU-70-0201 | SRR27223771 | New sequence | IV |
| IL-RIPHL-CAU-70-0202 | SRR27223716 | New sequence | IV |
| IL-RIPHL-CAU-70-0203 | SRR27223715 | New sequence | IV |
| IL-RIPHL-CAU-70-0204 | SRR27223714 | New sequence | IV |
| IL-RIPHL-CAU-70-0205 | SRR27223712 | New sequence | IV |
| IL-RIPHL-CAU-70-0206 | SRR27223711 | New sequence | IV |
| IL-RIPHL-CAU-70-0207 | SRR27223710 | New sequence | IV |
| IL-RIPHL-CAU-70-0208 | SRR27223659 | New sequence | IV |
| IL-RIPHL-CAU-70-0209 | SRR27223709 | New sequence | IV |
| IL-RIPHL-CAU-70-0210 | SRR27223708 | New sequence | IV |
| IL-RIPHL-CAU-70-0211 | SRR27223707 | New sequence | IV |
| IL-RIPHL-CAU-70-0212 | SRR27223706 | New sequence | IV |
| IL-RIPHL-CAU-70-0213 | SRR27223705 | New sequence | IV |
| IL-RIPHL-CAU-70-0214 | SRR27223704 | New sequence | IV |
| IL-RIPHL-CAU-70-0215 | SRR27223703 | New sequence | IV |
| IL-RIPHL-CAU-70-0216 | SRR27223701 | New sequence | IV |
| IL-RIPHL-CAU-70-0217 | SRR27223700 | New sequence | IV |
| IL-RIPHL-CAU-70-0218 | SRR27223699 | New sequence | IV |
| IL-RIPHL-CAU-70-0219 | SRR27223698 | New sequence | IV |
| IL-RIPHL-CAU-70-0220 | SRR27223697 | New sequence | IV |
| IL-RIPHL-CAU-70-0221 | SRR27223696 | New sequence | IV |
| IL-RIPHL-CAU-70-0222 | SRR27223695 | New sequence | IV |
| IL-RIPHL-CAU-70-0223 | SRR27223694 | New sequence | IV |
| IL-RIPHL-CAU-70-0224 | SRR27223693 | New sequence | IV |
| IL-RIPHL-CAU-70-0225 | SRR27223692 | New sequence | IV |
| IL-RIPHL-CAU-70-0226 | SRR27223690 | New sequence | IV |
| IL-RIPHL-CAU-70-0227 | SRR27223689 | New sequence | IV |
| IL-RIPHL-CAU-70-0228 | SRR27223688 | New sequence | IV |
| IL-RIPHL-CAU-70-0229 | SRR27223687 | New sequence | IV |
| IL-RIPHL-CAU-70-0230 | SRR27223686 | New sequence | IV |
| IL-RIPHL-CAU-70-0231 | SRR27223685 | New sequence | IV |
| IL-RIPHL-CAU-70-0232 | SRR27223652 | New sequence | IV |
| IL-RIPHL-CAU-70-0233 | SRR27223651 | New sequence | IV |
| IL-RIPHL-CAU-70-0234 | SRR27223650 | New sequence | IV |
| IL-RIPHL-CAU-70-0235 | SRR27223649 | New sequence | IV |
| IL-RIPHL-CAU-70-0236 | SRR27223647 | New sequence | IV |
| IL-RIPHL-CAU-70-0237 | SRR27223646 | New sequence | IV |
| IL-RIPHL-CAU-70-0238 | SRR27223645 | New sequence | IV |
| IL-RIPHL-CAU-70-0239 | SRR27223644 | New sequence | IV |
| IL-RIPHL-CAU-70-0240 | SRR27223643 | New sequence | IV |
| IL-RIPHL-CAU-70-0241 | SRR27223642 | New sequence | IV |
| IL-RIPHL-CAU-70-0242 | SRR27223641 | New sequence | IV |
| IL-RIPHL-CAU-70-0243 | SRR27223640 | New sequence | IV |
| IL-RIPHL-CAU-70-0244 | SRR27223639 | New sequence | IV |
| IL-RIPHL-CAU-70-0245 | SRR27223638 | New sequence | IV |
| IL-RIPHL-CAU-70-0246 | SRR27223636 | New sequence | IV |
| IL-RIPHL-CAU-70-0247 | SRR27223635 | New sequence | IV |
| IL-RIPHL-CAU-70-0248 | SRR27223634 | New sequence | IV |
| IL-RIPHL-CAU-70-0249 | SRR27223633 | New sequence | IV |
| IL-RIPHL-CAU-70-0250 | SRR27223632 | New sequence | IV |
| IL-RIPHL-CAU-70-0251 | SRR27223631 | New sequence | IV |
| IL-RIPHL-CAU-70-0252 | SRR27223630 | New sequence | IV |
| IL-RIPHL-CAU-70-0253 | SRR27223629 | New sequence | IV |
| IL-RIPHL-CAU-70-0254 | SRR27223628 | New sequence | IV |
| IL-RIPHL-CAU-70-0255 | SRR27223627 | New sequence | IV |
| IL-RIPHL-CAU-70-0256 | SRR27223625 | New sequence | IV |
| IL-RIPHL-CAU-70-0257 | SRR27223624 | New sequence | IV |
| IL-RIPHL-CAU-70-0258 | SRR27223623 | New sequence | IV |
| IL-RIPHL-CAU-70-0259 | SRR27223622 | New sequence | IV |
| IL-RIPHL-CAU-70-0260 | SRR27223621 | New sequence | IV |
| IL-RIPHL-CAU-70-0261 | SRR27223588 | New sequence | IV |
| IL-RIPHL-CAU-70-0262 | SRR27223587 | New sequence | IV |
| IL-RIPHL-CAU-70-0263 | SRR27223586 | New sequence | IV |
| IL-RIPHL-CAU-70-0264 | SRR27223585 | New sequence | IV |
| IL-RIPHL-CAU-70-0265 | SRR27223584 | New sequence | IV |
| IL-RIPHL-CAU-70-0266 | SRR27223582 | New sequence | IV |
| IL-RIPHL-CAU-70-0267 | SRR27223581 | New sequence | IV |
| IL-RIPHL-CAU-70-0268 | SRR27223580 | New sequence | IV |
| IL-RIPHL-CAU-70-0269 | SRR27223579 | New sequence | IV |
| IL-RIPHL-CAU-70-0270 | SRR27223578 | New sequence | IV |
| IL-RIPHL-CAU-70-0271 | SRR27223577 | New sequence | IV |
| IL-RIPHL-CAU-70-0272 | SRR27223576 | New sequence | IV |
| IL-RIPHL-CAU-70-0273 | SRR27223575 | New sequence | IV |
| IL-RIPHL-CAU-70-0274 | SRR27223574 | New sequence | IV |
| IL-RIPHL-CAU-70-0275 | SRR27223573 | New sequence | IV |
| IL-RIPHL-CAU-70-0276 | SRR27223570 | New sequence | IV |
| IL-RIPHL-CAU-70-0277 | SRR27223569 | New sequence | IV |
| IL-RIPHL-CAU-70-0278 | SRR27223568 | New sequence | IV |
| IL-RIPHL-CAU-70-0279 | SRR27223567 | New sequence | IV |
| IL-RIPHL-CAU-70-0280 | SRR27223566 | New sequence | IV |
| IL-RIPHL-CAU-70-0281 | SRR27223565 | New sequence | IV |
| IL-RIPHL-CAU-70-0282 | SRR27223564 | New sequence | IV |
| IL-RIPHL-CAU-70-0283 | SRR27223563 | New sequence | IV |
| IL-RIPHL-CAU-70-0284 | SRR27223562 | New sequence | IV |
| IL-RIPHL-CAU-70-0285 | SRR27223561 | New sequence | IV |
| IL-RIPHL-CAU-70-0286 | SRR27223559 | New sequence | IV |
| IL-RIPHL-CAU-70-0287 | SRR27223558 | New sequence | IV |
| IL-RIPHL-CAU-70-0288 | SRR27223557 | New sequence | IV |
| IL-RIPHL-CAU-70-0289 | SRR27223770 | New sequence | IV |
| IL-RIPHL-CAU-70-0290 | SRR27223769 | New sequence | IV |
| IL-RIPHL-CAU-70-0291 | SRR27223768 | New sequence | IV |
| IL-RIPHL-CAU-70-0292 | SRR27223767 | New sequence | IV |
| IL-RIPHL-CAU-70-0293 | SRR27223766 | New sequence | IV |
| IL-RIPHL-CAU-70-0294 | SRR27223765 | New sequence | IV |
| IL-RIPHL-CAU-70-0295 | SRR27223764 | New sequence | IV |
| IL-RIPHL-CAU-70-0296 | SRR27223762 | New sequence | IV |
| IL-RIPHL-CAU-70-0297 | SRR27223761 | New sequence | IV |
| IL-RIPHL-CAU-70-0298 | SRR27223760 | New sequence | IV |
| IL-RIPHL-CAU-70-0299 | SRR27223759 | New sequence | IV |
| IL-RIPHL-CAU-70-0300 | SRR27223758 | New sequence | IV |
| IL-RIPHL-CAU-70-0301 | SRR27223757 | New sequence | IV |
| IL-RIPHL-CAU-70-0302 | SRR27223756 | New sequence | IV |
| IL-RIPHL-CAU-70-0303 | SRR27223755 | New sequence | IV |
| IL-RIPHL-CAU-70-0305 | SRR27223754 | New sequence | IV |
| IL-RIPHL-CAU-70-0305 | SRR27223754 | New sequence | IV |
| IL-RIPHL-CAU-70-0306 | SRR27223753 | New sequence | IV |
| IL-RIPHL-CAU-70-0307 | SRR27223751 | New sequence | IV |
| IL-RIPHL-CAU-70-0308 | SRR27223750 | New sequence | IV |
| IL-RIPHL-CAU-70-0309 | SRR27223749 | New sequence | IV |
| IL-RIPHL-CAU-70-0310 | SRR27223748 | New sequence | IV |
| IL-RIPHL-CAU-70-0311 | SRR27223657 | New sequence | IV |
| IL-RIPHL-CAU-70-0312 | SRR27223747 | New sequence | IV |
| IL-RIPHL-CAU-70-0314 | SRR27223746 | New sequence | IV |
| IL-RIPHL-CAU-70-0315 | SRR27223745 | New sequence | IV |
| IL-RIPHL-CAU-70-0316 | SRR27223744 | New sequence | IV |
| IL-RIPHL-CAU-70-0317 | SRR27223743 | New sequence | IV |
| IL-RIPHL-CAU-70-0318 | SRR27223742 | New sequence | IV |
| IL-RIPHL-CAU-70-0319 | SRR27223740 | New sequence | IV |
| IL-RIPHL-CAU-70-0320 | SRR27223656 | New sequence | IV |
| IL-RIPHL-CAU-70-0321 | SRR27223655 | New sequence | IV |
| IL-RIPHL-CAU-70-0322 | SRR27223739 | New sequence | IV |
| IL-RIPHL-CAU-70-0323 | SRR27223684 | New sequence | IV |
| IL-RIPHL-CAU-70-0324 | SRR27223683 | New sequence | IV |
| IL-RIPHL-CAU-70-0325 | SRR27223682 | New sequence | IV |
| IL-RIPHL-CAU-70-0326 | SRR27223681 | New sequence | IV |
| IL-RIPHL-CAU-70-0327 | SRR27223653 | New sequence | IV |
| IL-RIPHL-CAU-70-0328 | SRR27223680 | New sequence | IV |
| IL-RIPHL-CAU-70-0329 | SRR27223679 | New sequence | IV |
| IL-RIPHL-CAU-70-0330 | SRR27223678 | New sequence | IV |
| IL-RIPHL-CAU-70-0331 | SRR27223677 | New sequence | IV |
| IL-RIPHL-CAU-70-0332 | SRR27223675 | New sequence | IV |
| IL-RIPHL-CAU-70-0333 | SRR27223674 | New sequence | IV |
| IL-RIPHL-CAU-70-0334 | SRR27223673 | New sequence | IV |
| IL-RIPHL-CAU-70-0335 | SRR27223672 | New sequence | IV |
| IL-RIPHL-CAU-038070-0583 | SRR29662443 | New sequence | III |
| IL-RIPHL-CAU-038070-0591 | SRR29662434 | New sequence | III |
| IL-RIPHL-CAU-038070-0593 | SRR29662432 | New sequence | III |
